# PheWAS and cross-disorder analyses reveal genetic architecture, pleiotropic loci and phenotypic correlations across 11 autoimmune disorders

**DOI:** 10.1101/2022.10.16.22281127

**Authors:** Apostolia Topaloudi, Pritesh Jain, Melanie B. Martinez, Josephine K. Bryant, Grace Reynolds, Petros Drineas, Peristera Paschou

## Abstract

Autoimmune diseases (ADs) are a group of more than 80 heterogeneous disorders that occur when there is a failure in the self-tolerance mechanisms triggering self-attacking autoantibodies. Most autoimmune disorders are polygenic and associated with genes in the human leukocyte antigen (HLA) region. However, additional non-HLA genes are also found to be associated with different ADs, and often these are also implicated in more than one disorder. Previous studies have observed associations between various health-related and lifestyle phenotypes and ADs. Polygenic risk scores (PRS) allow the calculation of an individual’s genetic liability to a phenotype and are estimated as the sum of the risk alleles weighted by their effect sizes in a genome-wide association study (GWAS). Here, for the first time, we conducted a comparative PRS-PheWAS analysis for 11 different ADs (Celiac Disease, Juvenile Idiopathic Arthritis, Multiple Sclerosis, Myasthenia Gravis, Primary Sclerosing Cholangitis, Psoriasis, Rheumatoid Arthritis, Systemic Lupus Erythematosus, Type 1 Diabetes, Vitiligo Early Onset, Vitiligo Late Onset) and 3,281 outcomes available in the UK Biobank that cover a wide range of lifestyle, socio-demographic and health-related phenotypes. We also explored the genetic relationships of the studied ADs, estimating their genetic correlation and performing cross-disorder GWAS meta-analyses for the identified AD clusters. In total, we observed 554 outcomes significantly associated with at least one disorder PRS, and 300 outcomes were significant after variants in the HLA region were excluded from the PRS calculations. Based on the genetic correlation and genetic factor analysis, we observed five genetic factors among studied ADs. Cross-disorder meta-analyses in each factor revealed genome-wide significant loci that are pleiotropic across multiple ADs. Overall, our analyses confirm the association of different factors with genetic risk for ADs and reveal novel observations that warrant further exploration.

## 1. Introduction

Autoimmune diseases (ADs) are a group of more than 80 (1) heterogeneous disorders that occur when there is a failure in the self-tolerance mechanisms triggering self-attacking autoantibodies(2). The estimated overall prevalence is 3% in the United States (3), and recent studies are reporting an increasing trend (4–6). Additionally, ADs are often comorbid and cluster within families (7,8). Most autoimmune disorders are polygenic and associated with genes in the human leukocyte antigen (HLA) region (9,10). However, many additional non-HLA genes are also found to be associated with different ADs, and many times they are implicated in more than one disorder (10). The genetic correlation across multiple ADs has not been fully explored (11,12). So far, cross-disorder GWAS meta-analyses have only focused on a few ADs, usually three to seven at a time (12–15), while others have only focused on pairwise meta-analyses (16,17). Given the wide comorbidities observed in epidemiological studies and the evidence for sharing of common genetic background across multiple ADs, a systematic large scale analysis is warranted.

In ADs, like other complex disorders, environmental factors are also involved in disease development along with genetic predisposition. Multiple studies have reported associations between viral infections and specific autoimmune diseases (18). For instance, a recent study (19) is suggesting that Epstein-Barr virus infection could be the leading cause of Multiple Sclerosis. Additional associations between ADs and environmental factors such as smoking and UV exposure have also been reported (20,21). Epidemiological studies have reported a high comorbidity across different ADs (8) as well as links to other traits, including psychotic disorders (22), allergies (23), and obesity (24).

Given the complex genetic background of ADs, Polygenic Risk Scores (PRS), allowing the calculation of an individual’s genetic liability to a phenotype, are an important tool to help understand disease correlations. They are usually estimated as the sum of the risk alleles weighted by their effect sizes in a genome-wide association study (GWAS) (25). This genetic risk can then become the basis of a Phenome-wide association study (PheWAS), with a goal to explore whether risk variants identified by a GWAS or disease PRS, are associated with a wide variety of phenotypes (26). Biobanks that combine genetic data with Electronic Health Records (EHR) are essential for the PheWAS approach, as they are the source of the phenotypes used in the analysis(27). Since the PheWAS is a hypothesis-free analysis, it can be used to generate new hypotheses about novel associations that might have not been uncovered through hypothesis-driven approaches.

Here, for the first time, we conducted a comparative PRS-PheWAS analysis for 11 different ADs and 3,281 outcomes available in the UK Biobank that cover a wide range of lifestyle, socio-demographic and health related phenotypes. Additionally, we explored the genetic relationships of the studied ADs, estimating their genetic correlation and performing cross-disorder GWAS meta-analyses for the identified AD sub-groups. Our findings provide an overview of the genetic and phenotypic architecture and relationships of ADs.

## 2. Methods

### 2.1. Study population

The UK Biobank is a large-scale, population-based, prospective cohort that recruited between 2006 and 2010 over 500,000 participants from the UK aged 40–69 years old. The participants provided blood, urine, and saliva samples for biochemical tests and genotyping, as well as self-reported information which was then linked to their health-related records. The phenotypic and genetic data we used in this study were obtained from UK Biobank under application number #61553.

The initial UK Biobank dataset included 488,377 individuals genotyped on the Affymetrix UK BiLEVE Axiom array or the Affymetrix UK Biobank Axiom array. We performed standard quality control on individuals and genetic markers (info>=0.9, maf>=0.01, geno<=0.02, hwe >= 10^−6^) with PLINK 1.9 (28). Initially, participants with withdrawn consent, sex mismatch, sex aneuploidy, self-reported non-white British ancestry, and with kinship coefficient <0.0625 (third-degree relatedness(29)) were excluded. Additional Principal Component Analysis (PCA) with 1000 Genomes data as reference was performed using TeraPCA (30) to exclude individuals with non-European ancestry. The final dataset included 330,841 individuals and 7,634,371 SNPs. 53.98% of the selected participants are females, the average age is 56.8 (sd=8) years. Table S1 provides a breakdown of the participants’ age and the percentage of the selected autoimmune diagnoses present in the UK Biobank.

### 2.2. PRS-pheWAS

#### 2.2.1. Polygenic Risk Scores

Publicly available and in-house GWAS summary statistics for 11 ADs performed on datasets of European ancestry and no UK Biobank participants were collected. For the PRS calculations we used PRSice2 (31). The independent SNPs with p-values<10^−5^, after clumping using a window of 500kb and an r^2^ threshold of 0.1, were included in the PRS calculations and the score was yielded as the weighted, standardized sum of the effect (score-std option). We repeated the PRS calculations excluding the extended HLA region (hg19, chr6 25-33 Mb). Table 1 shows the studied autoimmune datasets and the number of SNPs included in the PRS calculations.

**Table 1:** Autoimmune Disease datasets used in this study. The number of SNPs in the PRS calculations corresponds to the independent SNPs with p<10^−5^.

| Autoimmune Disorder | Abbreviation | Cases | Controls | SNPs in sumstats | SNPs in PRS | SNPs in PRS (no HLA) | PMID |
| --- | --- | --- | --- | --- | --- | --- | --- |
| Rheumatoid Arthritis | RA | 14,361 | 43,923 | 8,747,962 | 309 | 132 | 24390342 |
| Systemic Lupus Erythematosus | SLE | 4,036 | 6,959 | 7,915,251 | 200 | 144 | 26502338 |
| Vitiligo Late Onset | VITL | 1,467 | 19,156 | 7,552,975 | 77 | 49 | 30674883 |
| Vitiligo Early Onset | VITE | 704 | 9,031 | 8,020,475 | 84 | 60 | 30674883 |
| Type 1 Diabetes | T1D | 9,358 | 15,705 | 6,621,966 | 236 | 198 | 32005708 |
| Primary Sclerosing Cholangitis | PSC | 2,871 | 12,019 | 7,891,602 | 157 | 50 | 27992413 |
| Psoriasis | PSO | 2,997 | 9,183 | 161,173 | 191 | 121 | 23143594 |
| Multiple Sclerosis | MS | 9,772 | 17,376 | 472,086 | 147 | 86 | 21833088 |
| Celiac Disease* | CEL | 12,041 | 12,228 | 139,553 | 122 | 100 | 22057235 |
| Juvenile Idiopathic Arthritis | JIA | 2,816 | 13,056 | 122,330 | 45 | 5 | 23603761 |
| Myasthenia Gravis** | MG | 1,401 | 3,508 | 5,755,778 | 21 | 14 | 34400559 |

\*We included the summary statistics only from the European ancestry individuals in this study.
\*\* For the PRS calculations we used the summary statistics after excluding the UK Biobank samples, while for the rest of the analyses we included the full dataset described in the study.

### 2.3. Phenotypes

We included 3,281 phenotypes from UK Biobank that were assigned to seven broad categories: Biomarkers, Cognition and Mental Health, Disease Diagnoses, Health and Medical History, Physical Measures, Lifestyle, and Sociodemographics. Specifically for the Disease Diagnoses category, we included only the ICD10 codes and used the R PheWAS tool (32) to map similar diagnoses into one phecode. The breakdown of data fields in each category is shown in the Supplementary Materials (Figures S1-2).

### 2.4. PheWAS

For the PheWAS analyses, we used the tool PHESANT (33) to test the association of each disease PRS with each UK Biobank outcome. Age, sex, the first 10 principal components, and the genotyping batch were included as covariates in all regression models. To account for multiple testing, we used the R function p.adjust to calculate the FDR adjusted p-values and set the significance threshold at p_FDR_<0.05.

### 2.5. Cross-Disorder GWAS Meta-analysis

Pairwise genetic correlation analyses were performed for all 11 ADs after removing the extended HLA region (hg19, chr6 25-33 Mb) using LDSC(34). Only SNPs present in the HapMap3 reference panel were included in analyses and we used precalculated LD scores from 1000 Genomes European data. Datasets with less than 200,000 SNPs overlap with the LDSC reference data or heritability z-score <1.5 (as defined in (35)), were excluded from downstream analyses, namely CEL, PSO, and JIA datasets were removed.

To further explore the architecture and correlations of the studied disorders, we performed exploratory factor analysis (EFA) on the genetic correlation matrix using the R tool GenomicSEM (36). We further used a confirmatory factor analysis (CFA) to validate our model. For groups of disorders within each of the factors, we performed a cross-disorder GWAS meta-analysis with ReACt (37) and corrected for sample overlap between the datasets. In order to identify potentially pleiotropic SNPs, in each meta-analysis we estimated the posterior probability (m-value) using METASOFT (38) to identify SNPs with high m-values (m-value>0.9) for all studies in the meta-analysis. Then, using the pleiotropic SNPs, we identified the LD independent regions (r^2^<0.1) from the index SNPs with p<5×10^−8^. We used the default LD clumping window (250kb) and mapped into the regions the genes located no more than 20kb away. As reference for the LD estimation, we used the European samples from 1000 Genomes. Additionally, we merged into one overlapping genomic regions using bedtools (39). All genes that mapped to the identified LD independent regions for each meta-analysis after clumping, were submitted to g:Profiler(40) to perform functional enrichment analysis for Gene Ontology terms (GO:BP, GO:CC, GO:MF, released 2021-12-15), Reactome (REAC, released 2022-1-3) and Kyoto Encyclopedia of Genes and Genomes (KEGG FTP, released 2021-12-27). For all experiments we performed the recommended multiple hypothesis correction (g:SCS) method with the significance threshold of p = 0.05. We repeated the analysis after excluding the electronic GO annotations (Inferred from Electronic Annotation [IEA]) to have higher confidence in the enrichment analysis.

## 3. Results

### 3.1. Individual disorder phewas

First, we investigated the potential association of AD genetic risk to other phenotypes, including socioeconomic factors, lifestyle, biomarkers, disease diagnoses, health history and mental health, performing PRS-pheWAS. We used the LD-independent SNPs with p<10^−5^ to calculate PRS for each of the studied ADs in each individual, and tested the association of the autoimmune PRS, with 3,281 phenotypes in 330,841 UK Biobank samples (Table S1). Analysis was adjusted for age, sex, the first 10 principal components, and genotyping batch. We found a large number of associations with each disorder which differed depending on whether the HLA region was included in the analysis (Figure 1, Tables 2, Figure S3 and Table S2-S3). For SLE PRS with HLA region included in the analysis, we found the highest number of associations to different phenotypes (n=274). On the other hand, analysis for SLE PRS without the HLA region included, was associated with only 42 phenotypes. Interestingly, for CEL, and RA, more PRS associations to phenotypes were actually found when the HLA region was excluded from the calculations. For Psoriasis, genetic risk was found associated with other phenotypes only when HLA was included in the genetic risk calculations (significant association with 86 phenotypes).

**Figure 1:**
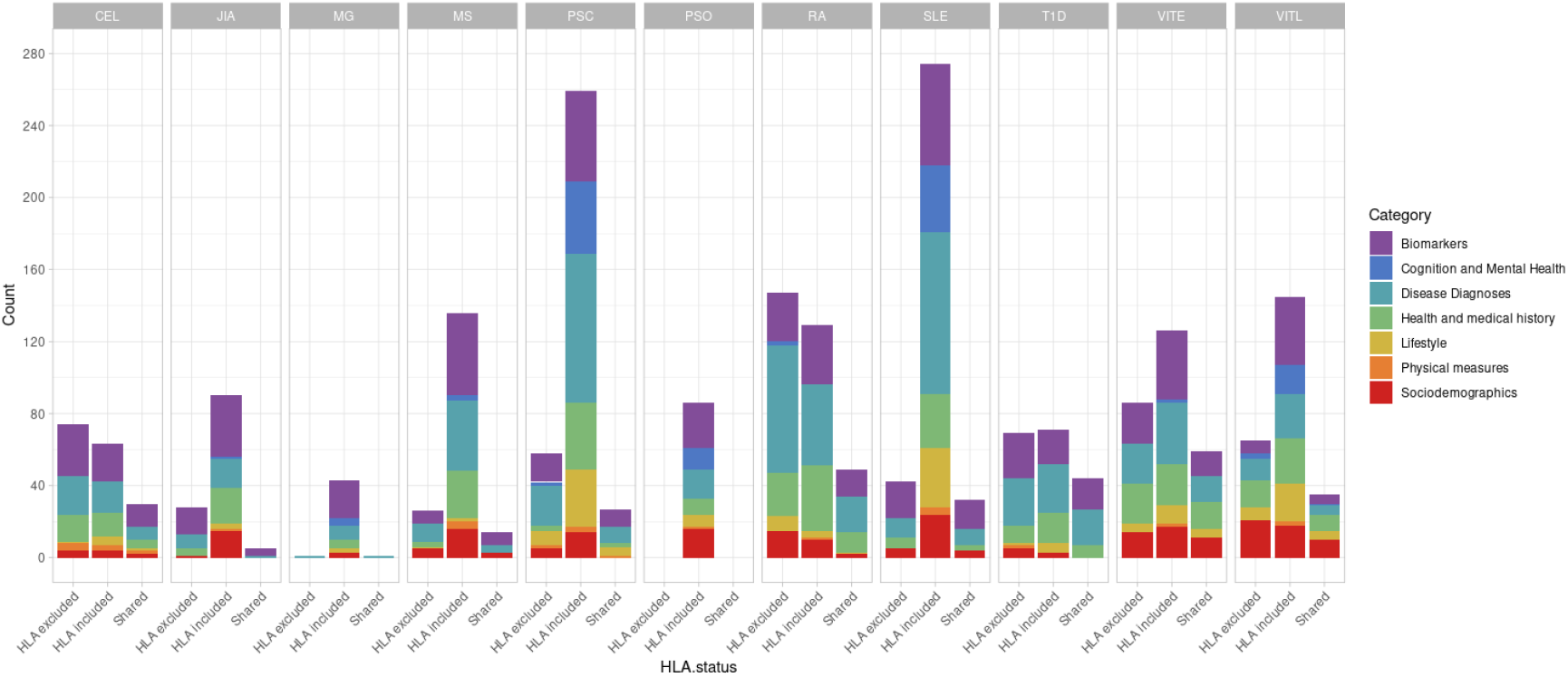
Distribution of phenotypes associated with autoimmune polygenic risk scores (p<10^−5^). The different colors represent general UK Biobank category. The “HLA excluded” bar shows the distribution of the significant associations with the phenotypes when HLA was excluded from the AD PRS calculations. The “HLA included” bar shows the distribution of the significant associations with the phenotypes when HLA was included in the AD PRS calculations. The “Shared” bar shows the distribution of the significant associations with the phenotypes for both HLA included or excluded AD PRSs.

**Table 2:** The most significant associations (p_FDR_<0.05) of each AD PRS and the UK Biobank phenotypes. The table shows the strongest associated phenotypes with each AD PRS with and without HLA, the beta, the 95% CI and the FDR adjusted p-value.

| Autoimmune Disorder | Top phenotype | HLA included |  |  | Top phenotype | HLA excluded |  |  |
| --- | --- | --- | --- | --- | --- | --- | --- | --- |
| | | $\beta$ | 95% Interval | Pfrd | | $\beta$ | 95% Interval | Pfrd |
| Celiac Disease | Celiac disease | 0.651 | [0.618 — 0.684] | <10-300 | Celiac disease | 0.539 | [0.497 — 0.581] | 7.13E-136 |
| Juvenile Idiopathic Arthritis | Celiac disease | -0.362 | [-0.406 — -0.319] | 1.26E-56 | Hypothyroidism NOS | 0.125 | [0.110 — 0.139] | 8.85E-57 |
| Multiple Sclerosis | Multiple sclerosis | 0.498 | [0.452 — 0.544] | 1.49E-97 | Multiple sclerosis | 0.406 | [0.353 — 0.460] | 2.85E-46 |
| Myasthenia Gravis | White blood cell (leukocyte) count | -0.03 | [-0.033 — -0.026] | 1.67E-59 | Hypothyroidism NOS | 0.036 | [0.021 — 0.051] | 9.22E-03 |
| Primary Sclerosing Cholangitis | Celiac disease | 0.699 | [0.667 — 0.731] | <10-300 | Eosinophill count | 0.03 | [0.027 — 0.034] | 1.37E-62 |
| Psoriasis | Psoriasis vulgaris | 0.371 | [0.336 — 0.407] | 9.66E-90 | *no significant outcome* |  |  |  |
| Rheumatoid Arthritis | Rheumatoid arthritis | 0.313 | [0.289 — 0.338] | 5.57E-132 | Hypothyroidism NOS | 0.204 | [0.189 — 0.218] | 1.69E-157 |
| Systemic Lupus Erythematosus | Celiac disease | 0.74 | [0.707 — 0.772] | <10-300 | Cystatin C | 0.021 | [0.018 — 0.024] | 3.71E-36 |
| Type 1 Diabetes | Type 1 diabetes | 0.324 | [0.287 — 0.360] | 1.31E-62 | Hypothyroidism NOS | 0.151 | [0.136 — 0.166] | 3.37E-82 |
| Vitiligo Early Onset | Skin colour | 0.173 | [0.165 — 0.180] | <10-300 | Skin colour | 0.257 | [0.249 — 0.264] | <10-300 |
| Vitiligo Early Onset | Ease of skin tanning | -0.14 | [-0.146 — -0.133] | <10-300 | Ease of skin tanning | -0.217 | [-0.223 — -0.211] | <10-300 |
| Vitiligo Late Onset | Skin colour | 0.151 | [0.143 — 0.159] | <10-300 | Skin colour | 0.267 | [0.259 — 0.274] | <10-300 |
| Vitiligo Late Onset | Ease of skin tanning | -0.125 | [-0.131 — -0.119] | <10-300 | Ease of skin tanning | -0.234 | [-0.240 — -0.227] | <10-300 |

In the following, we describe in detail patterns that emerge across all studied disorders and highlight significant results for phenotype associations to genetic risk with at least three ADs.

#### 3.1.1. Disease Diagnoses

For six of the studied ADs (CEL, RA, MS, SLE, T1D, VITE), we observed a significant positive association of PRS to the same disease diagnosis (Table S3). These results indicate a good predictive power of the respective PRS. We should note that for PSC, the disease diagnosis phenotype was not available in the dataset.

Celiac disease was found significantly associated with genetic risk for all 11 ADs that we studied. We observed that higher PRS for RA, VITE, VITL, JIA and PSO is associated with lower risk for the “Celiac disease” diagnosis phenotype. On the other hand, higher PRS for MS, MG, PSC, SLE, T1D and CEL was associated with higher risk with the “Celiac disease” diagnosis in the UK Biobank. The association with CEL, T1D, PSC, RA and JIA remained significant even after excluding the HLA, although with an opposite effect direction for the last two (Figure 2, Table S2).

**Figure 2:**
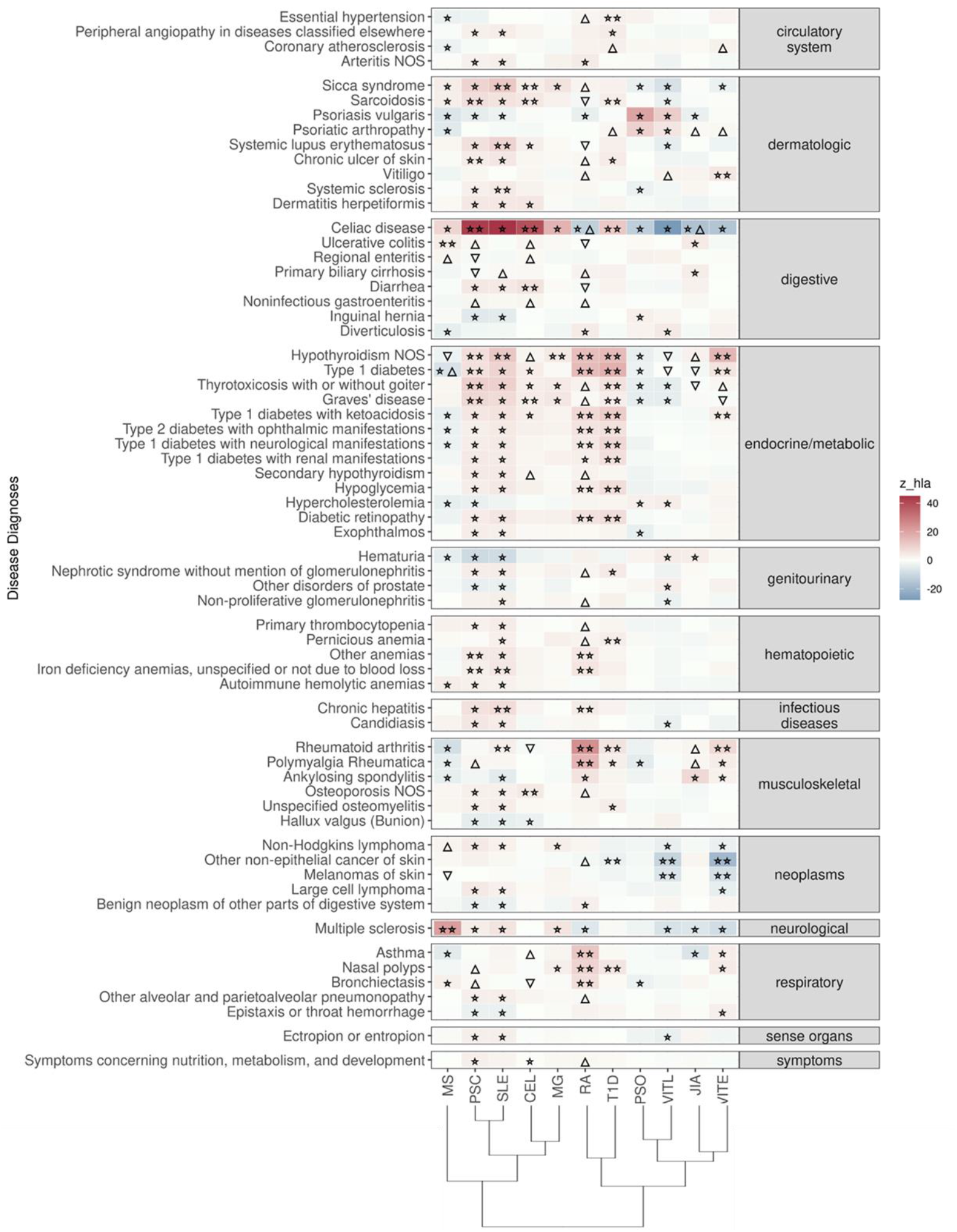
Significant PRS-PheWAS for at least three AD PRS with phenotypes in the Disease Diagnoses UK Biobank category. The shown phenotypes were significantly associated, after FDR adjustment, with at least three AD PRS irrespectively of the HLA status. The colors of cells indicate the standardized effect sizes (*β*) for the regression between AD PRS with HLA and each phenotype. The one star “☆” shows the significant results only with the “HLA included” AD PRS. The two stars “☆☆” show the significant associations with both “HLA included or excluded” AD PRS with the same effect direction. The star and the upper facing triangle “☆▵” show the significant associations with both “HLA included or excluded” AD PRS but with opposite effect directions. The upper facing triangle “▵” shows the significant associations only with “HLA excluded” AD PRS that the effect direction is the same as the color indicates. The down-facing triangle “▿” shows the significant associations only with “HLA excluded” AD PRS that the effect direction is the opposite of what the color indicates. For the grouping of the disease diagnoses phenotypes, we used the R PheWAS tool and collapse similar ICD-10 codes into one phecode. We used the hclust R function to perform the hierarchical clustering of the autoimmune disorders showing in the dendrogram using all standardized effect sizes for the disease diagnoses phenotypes.

“Ulcerative colitis” diagnosis was the second digestive phenotype most commonly found associated with the autoimmune PRS, and high MS, RA_(no-HLA)_, JIA_(HLA)_, PSC_(no-HLA),_ and CEL_(no-HLA)_ PRS were associated with higher risk for the diagnosis (Figure 2, Table S2).

In the endocrine diagnoses, most autoimmune PRS were associated with “Hypothyroidism” followed by “Type 1 diabetes”. RA, VITE, PSC, SLE, T1D and MG were associated with higher risk of “Hypothyroidism”, even after HLA was excluded. JIA, VITL, MS and CEL association with Hypothyroidism was significant only after HLA was excluded (Figure 2, Table S2).

In dermatologic diagnoses, the autoimmune “Sicca syndrome” was the most associated phenotype with the autoimmune PRS. We observed a positive association of the “Sicca syndrome” diagnosis with SLE, CEL, RA_(no-HLA)_, MS_(HLA)_, PSC_(HLA)_, and MG_(HLA)_ PRS, whereas there was a negative association with PRS VITE, PSO, and VITL (Figure 2, Table S2).

In the neoplasms category, high PRS for VITL and VITE was associated with lower risk for skin cancer outcomes, including Non-Hodgkins lymphoma, other non-epithelial cancer of skin and melanomas of skin (Figure 2, Table S2).

#### 3.1.2. Cognition and mental health

For PSC and SLE PRS, we found the largest overlap (n=20) of traits associated in the same direction. These associations included lower risk for phenotypes such as addictions, depression, and “low/worse” mental health, while they were positively associated with phenotypes describing higher cognitive function (Figure 3, Table S2). For PRS of MS, and MG, we also found an association with lower risk for phenotypes describing poor mental health (Figure 3, Table S2). On the contrary, higher PRS for VITL was associated with phenotypes describing poor mental health and depression (n=10), and had a negative association with phenotypes describing cognitive function (n=4) (Figure 3, Table S2). PSO and VITE associated with higher risk with phenotypes describing poor mental health and anxiety (Figure 3, Table S2).

**Figure 3:**
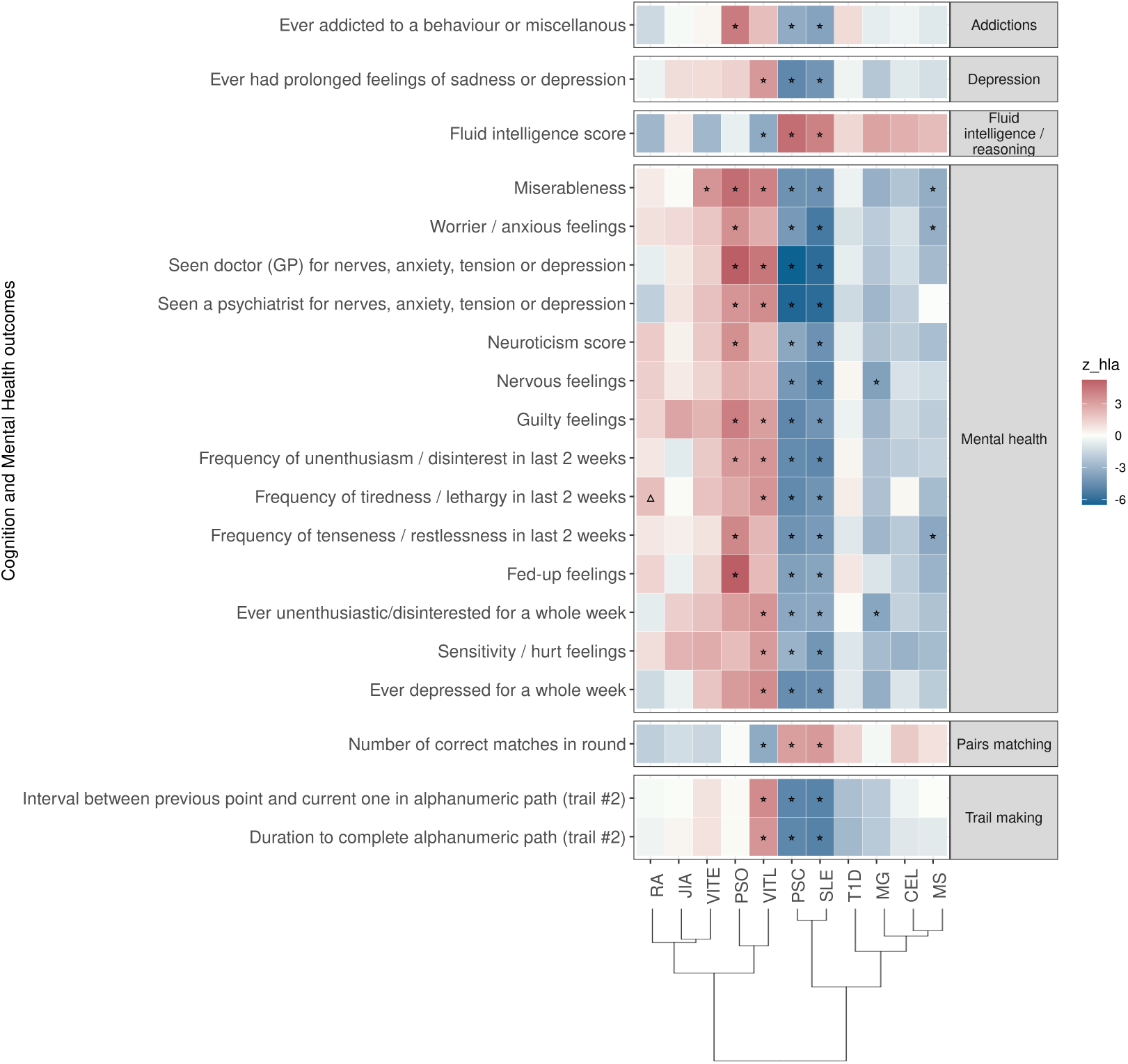
Significant PRS-PheWAS for at least three AD PRS with phenotypes in the Cognition and Mental Health UK Biobank category. The shown phenotypes were significantly associated, after FDR adjustment, with at least three AD PRS irrespectively of the HLA status. The colors of cells indicate the standardized effect sizes (*β*) for the regression between AD PRS with HLA and each phenotype. The one star “☆” shows the significant results only with the “HLA included” AD PRS. The upper facing triangle “▵” shows the significant associations only with “HLA excluded” AD PRS that the effect direction is the same as the color indicates. For the grouping of the phenotypes, we used the categories provided by the UK Biobank. We used the hclust R function to perform the hierarchical clustering of the autoimmune disorders showing in the dendrogram using all standardized effect sizes for the Cognition and Mental Health phenotypes.

#### 3.1.3. Lifestyle

In this category the trait “Never eat eggs, dairy, wheat, sugar: Wheat products” was associated with PRS for ten ADs when HLA was included in analysis, suggesting susceptibility to food allergies; VITL, VITE, PSO, JIA, RA are negatively associated with the phenotype, while PSC, SLE, MG, CEL (irrespectively of HLA) and T1D were positively associated with the phenotype (Figure 4, Table S2).

**Figure 4:**
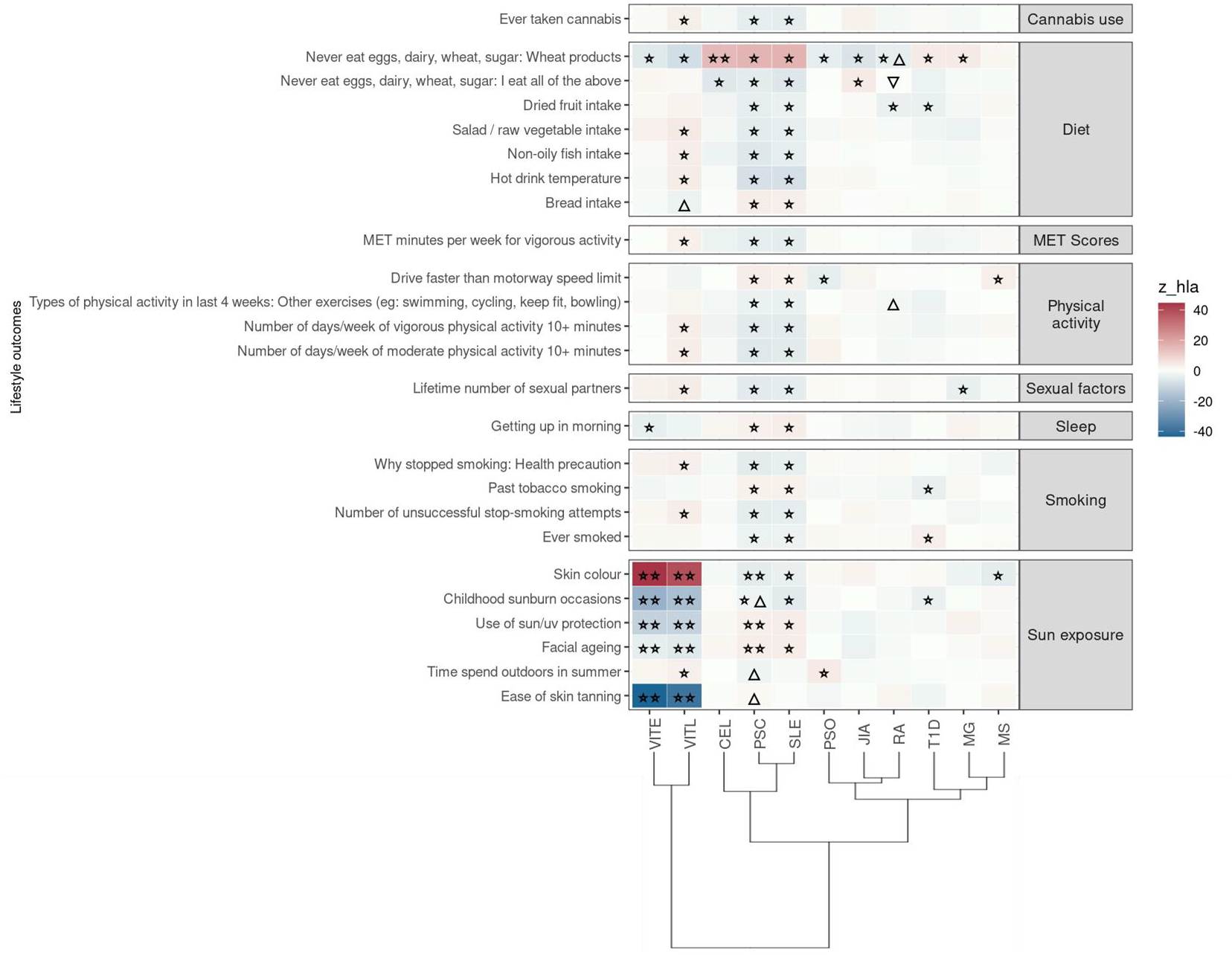
Significant PRS-PheWAS for at least three AD PRS with phenotypes in the Lifestyle UK Biobank category. The shown phenotypes were significantly associated, after FDR adjustment, with at least three AD PRS irrespectively of the HLA status. The colors of cells indicate the standardized effect sizes (*β*) for the regression between AD PRS with HLA and each phenotype. The one star “☆” shows the significant results only with the “HLA included” AD PRS. The two stars “☆☆” show the significant associations with both “HLA included or excluded” AD PRS with the same effect direction. The star and the upper facing triangle “☆▵” show the significant associations with both “HLA included or excluded” AD PRS but with opposite effect directions. The upper facing triangle “▵” shows the significant associations only with “HLA excluded” AD PRS that the effect direction is the same as the color indicates. For the grouping of the phenotypes, we used the categories provided by the UK Biobank. We used the hclust R function to perform the hierarchical clustering of the autoimmune disorders showing in the dendrogram using all standardized effect sizes for the Lifestyle category phenotypes.

Again, same as for the previous category of traits, PSC and SLE PRS had the largest overlap of associated phenotypes (n=23) in the same effect direction. They were negatively associated with phenotypes related to dietary habits (higher intake of dried fruit, salad/ raw vegetable, non-oily fish), cannabis usage, exercise, smoking status (Figure 4, Table S2).

Additionally, VITL and VITE PRS (irrespectively of HLA) were positively associated with darker skin color, and negatively associated with higher risk of “ease of skin tanning”, “childhood sunburn occasions”, “use of sun/uv protection” and “facial aging” (Figure 4, Table S2). We observed the opposite associations between PSC and SLE PRS and these sun exposure phenotypes, except for the “childhood sunburn occasions” (Figure 4, Table S2).

#### 3.1.4. Health and medical history

In this category the self-reported phenotype “Diagnosed with coeliac disease or gluten sensitivity” was significantly associated with 11 autoimmune PRS (Figure S4, Table S2). We observed a positive association with PSC, SLE, T1D, CEL, MG, and a negative association with RA, JIA, VITL, VITE, PSO, these results are consistent with similar phenotypes, such as the “Celiac disease” diagnosis and the “Never eat eggs, dairy, wheat, sugar: Wheat products” phenotype in the Lifestyle category.

Additionally, high PRS T1D, VITL, and VITE were associated with lower risk for “Basal cell carcinoma” phenotype under the Cancer register sub-category. Specifically for VITE and VITL we observed a negative association with self-reported basal cell carcinoma (Figure S4, Table S2).

#### 3.1.5. Sociodemographics

In this category, we observed a positive association of PSO_(HLA)_ PRS with Indices of Multiple Deprivation, such as Health, Employment, Income, and Education scores, as well as the Index of Multiple Deprivation (Figure S5, Table S2). RA_(no-HLA)_ PRS was also positively associated with Health and Employment Deprivation Indices. These results indicate that the participants with higher PSO and RA PRS had a higher deprivation index and lower levels of education, health, income and employment. Whereas VITE was associated with lower risk for Health, Employment, and Education, Deprivation scores, as well as the Index of Multiple Deprivation. Only when the HLA was excluded from the VITE PRS calculation, we observed a negative association with the Income Deprivation score. VITL_(no-HLA)_ PRS was negatively associated with Health, Employment, Income, and Education, Deprivation scores, as well as the Index of Multiple Deprivation, which indicates the participants with higher VITE and VITL PRS, had higher levels education, health, income and employment.

We also observed significant associations of autoimmune PRS with phenotypes in the Biomarkers (Figure S6-S8) and Physical Measures (Figure S9) categories, without any patterns emerging across disorders. Results are shown in supplement.

### 3.2. Cross-Disorder GWAS Meta-analysis

Driven by the known comorbidity across AD (based on epidemiological studies (8) and the overlap in phenotypic associations with autoimmune PRS that we described above, we proceeded to perform cross-disorder genetic correlation and GWAS summary statistics meta-analyses to explore the genetic relationship and genetic architecture of ADs and identify potentially pleiotropic loci. Such pleiotropic loci would drive pathophysiology across multiple ADs.

Initially, we performed analysis for all 11 ADs (Figure S10), however, given the limited SNP overlap of our datasets for CEL, PSO and JIA we excluded them from further analyses. After correction for multiple testing, we observed significant positive correlations of RA with T1D (rg_no-HLA_:0.52), SLE (rg_no-HLA_:0.51), and MG (rg_no-HLA_:0.47). VITL and VITE were also significantly correlated (rg_no-HLA_:0.64). Additional autoimmune correlations with p<0.05 are shown in Figure 5A-B, including the pairwise correlations after excluding HLA.

**Figure 5:**
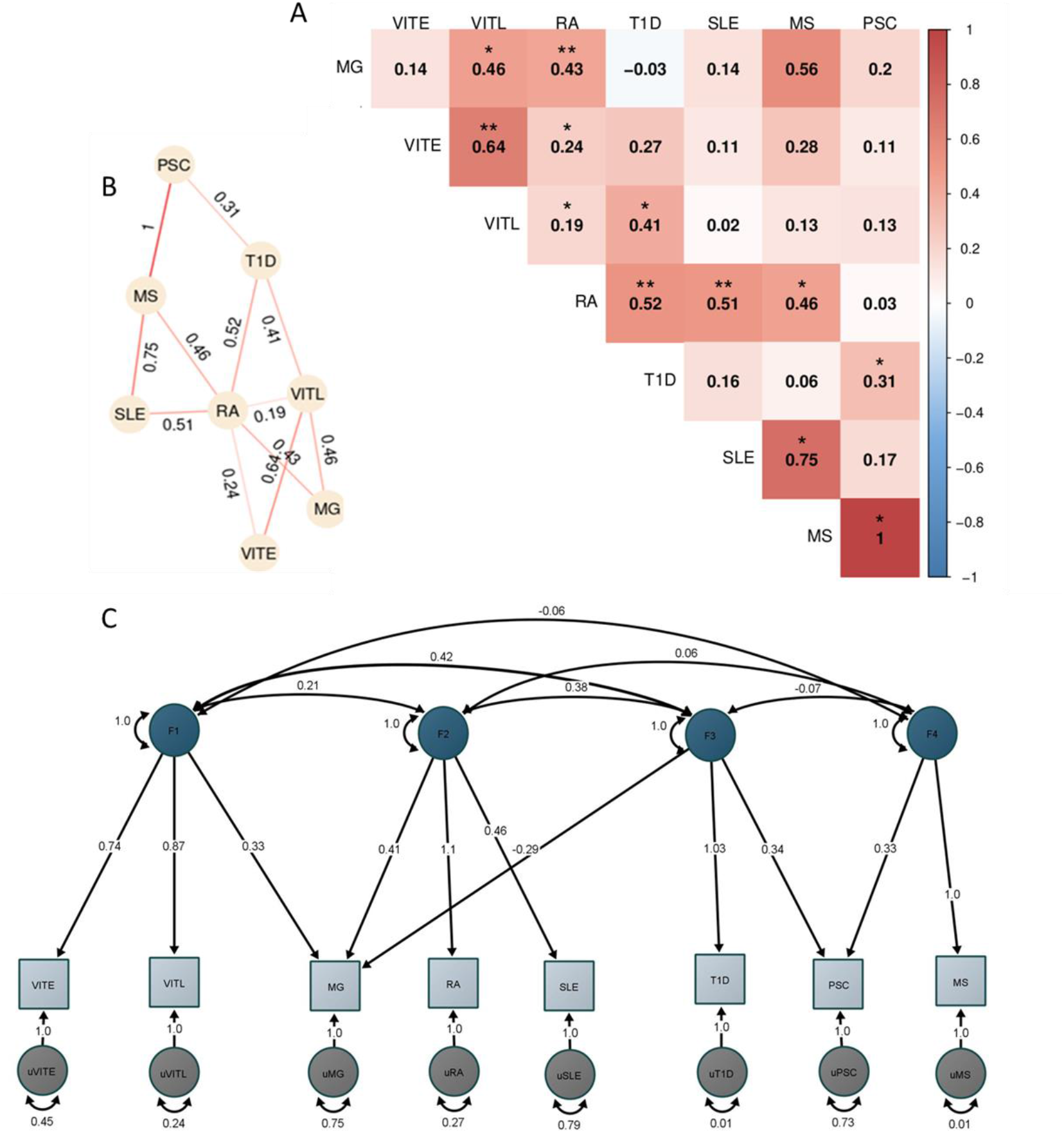
Genetic correlation and factor analysis for 8 autoimmune disorders. The figure shows the analyses of the 8 autoimmune disorders with enough overlap (>200.000 SNPs) with HapMap3 data provided by LDSC after excluding the HLA locus (hg19, chr6 25-33 Mb). A) Heatmap of the pairwise LDSC genome wide genetic correlations of the 8 autoimmune disorders after excluding the SNPs in the HLA region. The red color reflects more positive correlation coefficients while blue reflects more negative coefficients, and the numbers within each cell are the correlation coefficients. The correlations with p<0.05 are denoted with one asterisk (*), while the two asterisks show the correlations that are significant after the Bonferroni correction. B) Network representation of the genetic correlation between the autoimmune disorders with p<0.05. The numbers show the correlation coefficient and the stronger the line color shows a higher coefficient. C) Path graph of the confirmatory factor model estimated using the Genomic SEM. Four factors were identified. The factor loadings for each trait are depicted by arrows between the trait and the factor, with the standardized loading value and the standard error in the parentheses. Correlation between factors is indicated by arrows between them. Residual variance for each trait is indicated by the two-headed arrow connecting the variable to itself.

Based on the fact that the pairwise genetic correlation analysis showed a complicated correlation pattern among the studied disorders, we performed exploratory factor analysis (EFA) followed by a confirmatory factor analysis (CFA) to dissect the AD relationships. We used the four-factor model identified in EFA and included the disorders with loadings greater than |0.3| in each factor. The CFA analysis in GenomicSEM showed a good fit of the model to the data (χ^2^(12) =16.1; AIC =64.1; CFI = 0.98; SRMR = 0.07). The first factor included VITE, VITL and MG. MG, RA and SLE were included in the second factor, while the third factor consisted of T1D, PSC and MG (with a negative loading). Lastly, factor four consisted of PSC and MS (Figure 5C).

In the cross-disorder meta-analysis on the first factor, that includes VITL, VITE and MG, we identified nine significant pleiotropic (m-value>0.9 in all studies) LD independent regions (Table 3); two of them mapping on the *Cytotoxic T-Lymphocyte Associated Protein 4* (CTLA4) -*Inducible T Cell Costimulator* (ICOS) and Fli-1 Proto-Oncogene, ETS Transcription Factor (FLI1) genes, were not significant in the individual GWAS studies included here. However in the GWAS catalog (41) FLI1 has a significant association with Vitiligo, when the onset age is not taken into account (42), and CTLA4 is found associated with different GWASs (not studied here) for both Myasthenia gravis (43) and Vitiligo (42). CTLA4 was also significant in the gene-based analysis (44) of the data we used in this meta-analysis. The gene set enrichment analysis including the genes in the significant and pleiotropic regions identified four significantly enriched GO:BP terms; bone cell development, immune system development, myeloid cell development, and immune system process (Figure 6A, Table S4).

**Table 3:** Genome-wide significant (p<5×10^−8^) LD independent loci from the VITL-VITE-MG meta-analysis. The column SNP contains the top SNP in each locus. The columns P, OR and SE correspond to the top SNP in each locus. The autoimmune disorder specific Top SNP column contains the top genome-wide significant SNP in the locus that was available in the input dataset. The autoimmune disorder specific P column contains the lowest p-value in the locus that was available in the input dataset.

| SNP | Locus | Nearest genes (<20kb) | P-meta | OR | SE | Top SNP VITE | P-VITE | Top SNP VITL | P-VITL | Top SNP MG | P-MG |
| --- | --- | --- | --- | --- | --- | --- | --- | --- | --- | --- | --- |
| rs9981704 | chr21:43831955-43867059 | TMPRSS3<br>UBASH3A | 6.09x10<sup>-19</sup> | 1.31 | 0.03 | - | 4.30x10<sup>-4</sup> | rs12482396 | 4.06x10<sup>-20</sup> | - | 3.60x10<sup>-3</sup> |
| rs60946162 | chr3:188084682-188133518 | LPP | 1.78x10<sup>-15</sup> | 0.8 | 0.03 | - | 2.43x10<sup>-7</sup> | rs13098877 | 2.62x10<sup>-11</sup> | - | 1.17x10<sup>-3</sup> |
| rs7137828 | chr12:111833788-112037526 | ATXN2<br>SH2B3 | 1.70x10<sup>-13</sup> | 0.82 | 0.03 | - | 7.31x10<sup>-5</sup> | rs10774624 | 8.28x10<sup>-9</sup> | - | 1.26x10<sup>-3</sup> |
| rs8088891 | chr18:60007263-60029292 | TNFRSF11A | 3.51x10<sup>-13</sup> | 1.22 | 0.03 | - | 9.89x10<sup>-4</sup> | - | 1.14x10<sup>-3</sup> | rs4369774 | 1.09x10<sup>-13</sup> |
| rs1951459 | chr6:167370353-167455629 | FGFR1OP<br>MIR3939<br>RNASET2 | 3.61x10<sup>-13</sup> | 0.81 | 0.03 | rs2247315 | 3.48x10<sup>-9</sup> | rs366938 | 1.14x10<sup>-8</sup> | - | 9.53x10<sup>-4</sup> |
| rs64547 | chr22:37575469-37595156 | C1QTNF6<br>SSTR3 | 8.14x10<sup>-13</sup> | 0.82 | 0.03 | rs229528 | 2.09x10<sup>-8</sup> | rs229527 | 1.55x10<sup>-12</sup> | - | 7.33x10<sup>-3</sup> |
| rs831071 | chr3:71426419-71430389 | FOXP1 | 1.01x10<sup>-10</sup> | 1.2 | 0.03 | - | 2.11x10<sup>-3</sup> | rs60135207 | 3.14x10<sup>-9</sup> | - | 3.40x10<sup>-6</sup> |
| rs3116513 | chr2:204694611-204792732 | CTLA4<br>ICOS | 1.42x10<sup>-10</sup> | 1.19 | 0.03 | - | 2.25x10<sup>-2</sup> | - | 5.64x10<sup>-5</sup> | - | 2.65x10<sup>-6</sup> |
| rs644515 | chr11:128589472-128617231 | FLI1 | 1.44x10<sup>-9</sup> | 1.17 | 0.03 | - | 2.89x10<sup>-5</sup> | - | 2.89x10<sup>-5</sup> | - | 1.24x10<sup>-2</sup> |

**Figure 6:**
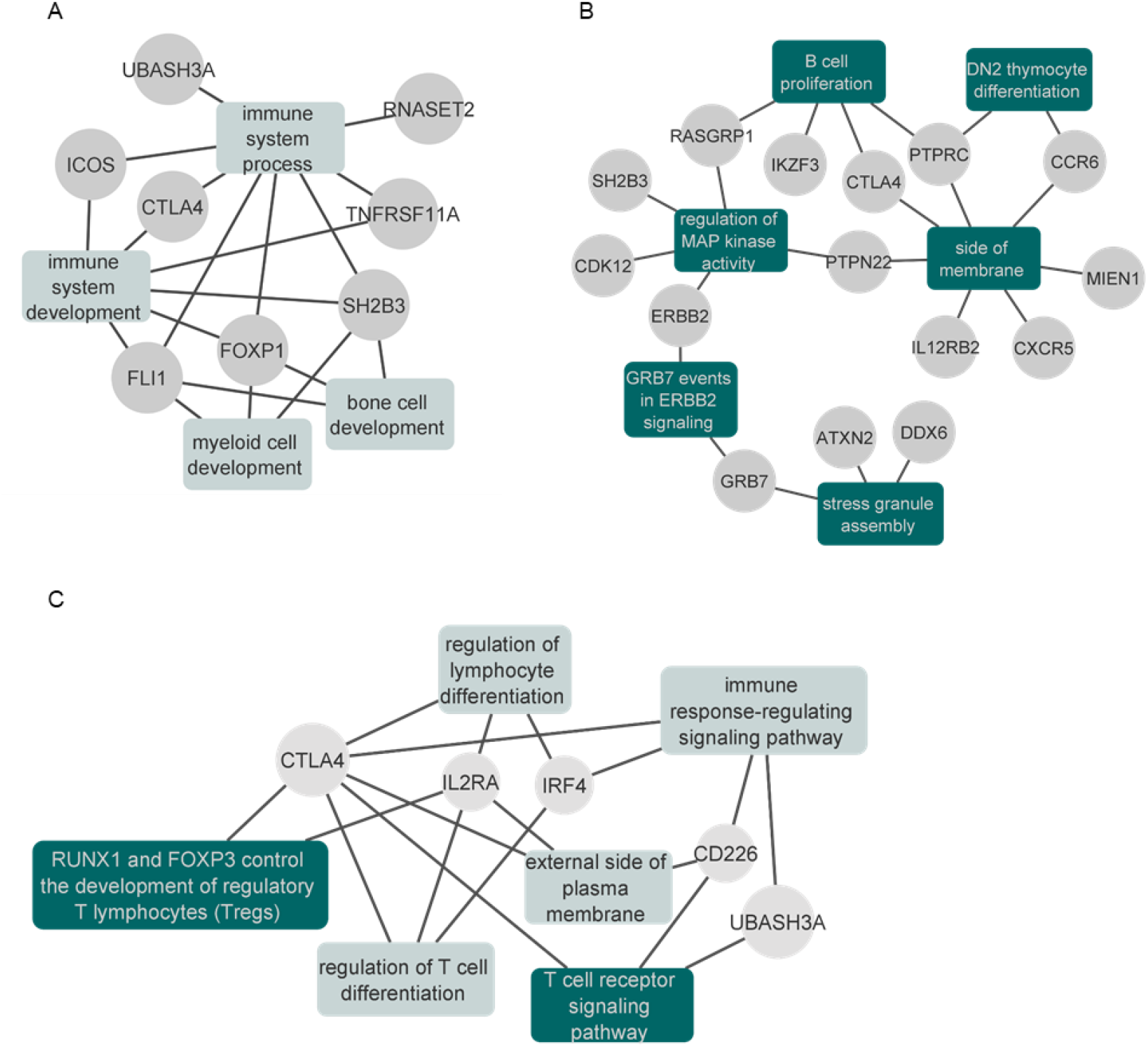
Network plots of the enrichment analysis for the cross-disorder meta-analyses. A) Results of the significantly enriched terms from the genes identified in the VITE-VITL-MG meta-analysis. Results are also shown in Supplementary Table 4. B) Results of the significantly enriched terms (after excluding the IEA terms) from the genes identified in the SLE-MG-RA meta-analysis. The full results are also shown in the Supplementary Table 6. C) Results of the significantly enriched terms from the genes identified in the T1D-MG-PSC meta-analysis. The full results are also shown in the Supplementary Table 8. Enriched gene sets that remained significant after excluding the IEA GO terms are shown in dark green.

When we performed the meta-analysis of MG, RA and SLE, we identified 17 genomewide significant pleiotropic loci. Three of these loci mapping to *Protein Tyrosine Phosphatase Receptor Type C* (PTPRC), *Interleukin 12 Receptor Subunit Beta 2* (IL12RB2) and LINC00824 were not genome-wide significant in the GWAS studies we analyzed (focusing on European ancestry), however, they were reported as significant associations in GWAS of higher power that were multi-ethnic (Table S5). The gene set enrichment analysis of genomewide significant and pleiotropic regions identified 22 significantly enriched terms. Among them, six (DN2 thymocyte differentiation, regulation of MAP kinase activity, stress granule assembly, B cell proliferation, side of membrane, GRB7 events in ERBB2 signaling) were significant even after excluding the IEA GO terms (see methods) (Figure 6B, Table S6).

In the meta-analysis of MG, T1D and PSC, we observed seven pleiotropic and genome wide significant loci. One of them, found on chr4:10,709,726-10,726,520 (closest gene *Cytokine Dependent Hematopoietic Cell Linker* (CLNK), 23Kb downstream) has not been previously found to be associated with either of the three disorders (Table S7). The gene-set enrichment analysis identified six significantly enriched terms after multiple testing correction, with “RUNX1 and FOXP3 control the development of regulatory T lymphocytes (Tregs)” and “T cell receptor signaling pathway” as the two top terms (Table S8). These two were the only significant terms when we repeated the analysis after excluding the IEA GO terms (see methods)(Figure 6C, Table S8).

Finally, for the cross-disorder meta-analysis of PSC and MS, we identified two genome-wide significant and pleiotropic loci, mapping to the previously associated *Interleukin 2 Receptor Subunit Alpha* (IL2RA) and *BTB Domain And CNC Homolog 2* (BACH2) genes (Table S9). The gene-set enrichment analysis identified three significant terms, primary adaptive immune response, primary adaptive immune response involving T cells and B cells, and interleukin-2 receptor complex. All of them remained significant even after we excluded the IEA GO terms (Table S10).

## 4. Discussion

We report results on the first PRS-PheWAS analysis exploring the association of genetic risk for 11 autoimmune disorders and 3,281 phenotypes on 330,841 individuals of European ancestry from the UK Biobank. Additionally, we explored the genetic relationship between the studied ADs seeking to dissect the genetic architecture of these highly correlated and often comorbid phenotypes.

We were able to recover previously identified associations between ADs based on epidemiological or genetic studies. For instance, a study in a Taiwanese population showed higher risk of first-degree relatives with Sicca to develop other autoimmune disorders including SLE, MS, MG, and RA (45); and we also observed here a positive association between the PRS of these four ADs and Sicca syndrome outcome. Other autoimmune-related diagnosis outcomes associated with higher risk for the studied ADs, included hypothyroidism and Graves disease. A link between those disorders and RA, Vitiligo, SLE, T1D, CEL, and MG, is also supported by the literature (46–50). Additionally, as reported in previous studies (51–53), we also observed a negative association between Vitiligo risk and skin cancer.

Interestingly, we observed many associations with environmental and lifestyle factors. Diet and specifically the consumption of non-wheat products was the outcome that we found to be associated with the risk for most of the studied ADs pointing to gluten intolerance and food allergies. We observed a significant positive association between PSC, SLE, CEL, MG, T1D, and RA (when HLA was excluded from PRS calculations) and not consuming wheat, while the association was negative for VITE, VITL, PSO, JIA and RA. We also observed the same pattern of AD associations with the Celiac disease diagnosis phenotype, which could be indicative of the connection between gluten intolerance and Celiac disease. There is prior evidence suggesting that a gluten-free diet could be beneficial not only for patients with Celiac disease but also for T1D, RA, MS, autoimmune hepatitis, and PSO (54).

Smoking was another factor that we found significantly associated with SLE and PSC genetic risk. Indeed, it has been previously suggested that smoking is associated with higher risk for double-stranded DNA seropositivity, a marker used for SLE diagnosis, in SLE patients (55), while for PSC, there is some evidence to suggest that smoking is associated with lower risk for developing the disease (56,57), although not always consistently supported (58). Interestingly, a previous study found that severe sunburn incidents and higher tanning ability in women are associated with higher risk of developing Vitiligo (59), however, we actually observed the opposite association for Vitiligo genetic risk, perhaps indicating different behavior towards sun exposure based on genetic risk. We also observed a negative association between CEL genetic risk and the weight of the first child. Previous studies have shown that women with undiagnosed or untreated celiac disease have higher risk to deliver a baby with reduced birthweight (60).

The link between autoimmune disorders and mental health has been previously described. For instance, exposure to stress-related disorders was found to be associated with higher risk for ADs (61), and both positive and negative associations of ADs with psychotic disorders have been summarized elsewhere (22). In our study, we observed that risk for VITL and PSO was positively associated with self-reported outcomes describing poor mental health, which is in line with previous works (22,61–63). We also observed that risk for SLE and PSC was associated with better mental health outcomes. Epidemiological studies have reported higher psychological distress in patients with SLE and PSC (61,64–67). However, in a study exploring the genetic correlation between immune and psychiatric related phenotypes using GWAS summary statistics, SLE was found to be significantly positively correlated only with Schizophrenia and no other psychiatric phenotypes (68). Additionally, a study using Mendelian Randomization between SLE and depression showed SLE genetic variants mildly reduce the odds of depression, suggesting that the observed association between SLE and depression might not be attributed to genetic factors (69). Thus, further analyses could be useful to explore the gap between the associations between SLE and mental health phenotypes observed in epidemiological studies, but not when using genetic data.

It is well demonstrated that ADs are often comorbid and share both HLA and non-HLA genetic loci (8–10,15). In a recent study (11), where the genetic correlation between 13 (7 of them are also studied here) autoimmune and inflammatory disorders was also explored, the authors observed correlations across ADs and similar patterns to what we also found. Furthermore, we also provide here a more detailed analysis to understand the genetic architecture of ADs including EFA to reveal subgroups of disorders and cross-disorder GWAS to reveal pleiotropic loci that could underlie multiple disorders and drive comorbidities. Indeed, in line with the existing notion of shared genetic background across ADs, we detected numerous genome-wide significant and pleiotropic loci in each meta-analysis. All except one had already been previously associated with at least one of the ADs included in the meta-analysis, or were associated with the traits in studies of different ancestries or larger sample GWAS which we could not analyze here because summary statistics data were not available. Importantly, we identify one novel genome-wide significant and pleiotropic locus in the meta-analysis of T1D-MG-PSC. This is a previously unknown locus that could play a role in the etiology of all three disorders and is found 23Kb downstream of *CLNK* gene that encodes Clnk, an adapter of the SLP76 family, is involved in the regulation of immunoreceptor signaling (70).

This study comes with both strengths and limitations. The PheWAS analysis allowed us to detect significant associations between AD risk and multiple phenotypes, even after excluding the HLA region. Additionally, we were able to detect pleiotropic loci in the autoimmune subgroups that are involved in immune-related processes as the gene-set enrichment analysis revealed. However, there are limitations in this study that should be taken into account when interpreting the results. For the PRS calculations, although we used the largest AD summary statistics data available, there were differences in power regarding their sample size and number of SNPs. Also, as the number of UK Biobank participants with AD diagnoses is limited, we were not able to calculate the optimal p-value threshold for SNPs to be included in PRS calculations, but rather set as threshold the p-value 10^−5^.

In conclusion, in this study we observed ADs PRS to be associated with multiple health-related and environmental factors, even after excluding the HLA region, and explored the genetic relationships of the selected ADs by estimating their genetic correlation and identifying pleiotropic genetic regions that underlie genetic risk across multiple ADs. Overall, our analyses indicate potential factors associated with genetic risk for ADs, some of which have been reported previously, and also novel observations that need to be further explored. These results are suggesting that the assessment of additional exposures related to lifestyle, mental and physical health risks by clinicians, could be beneficial for individuals with higher risk for autoimmune disorders.

## Data Availability

All data produced in the present work are contained in the manuscript

## Supporting Information

**Table S1 Demographic information of the 330**,**841 UK Biobank participants included in the analysis**.

The ICD10 Disease Diagnoses category includes the diagnoses for the autoimmune disorders that are included in this study and were present in the UK Biobank.

**Table S2 PRS-pheWAS results for association of genetic risk of 11 autoimmune disorders with 3**,**281 phenotypes in UK Biobank**.

Genetic risk scores were calculated as the weighted standardized sum of the effect of independent SNPs with p-values<10^−5^ for each disorder. The estimation of the genetic risk scores was repeated after excluding the extended HLA region (hg19, chr6 25-33 Mb). Estimates were generated by PHESANT.

**Table S3: Associations of AD PRS with disease diagnosis phenotype for the same disorder**. The table shows the associations of the disease diagnosis phenotypes and the same AD PRS with and without HLA, only for the ADs that the same diagnosis was available. For the Vitiligo early and late onset we used the general Vitiligo diagnosis phenotype that was available in the UK Biobank.

**Table S4 Significantly enriched gene sets genes identified in the VITL-VITE-MG meta-analysis**.

The p.Val column is the adjusted p-value using the suggested g:SCS method by the g:Prolifer tool used for the analysis. The Genes column contains the provided genes that are in each identified gene set.

**Table S5 Genome-wide significant (p<5×10-8) LD independent loci from the SLE-RA-MG meta-analysis**.

The column SNP contains the top SNP in each locus. The columns P, OR and SE correspond to the top SNP in each locus. The autoimmune disorder specific Top SNP column contains the top genome-wide significant SNP in the locus that was available in the input dataset. The autoimmune disorder specific P column contains the lowest p-value in the locus that was available in the input dataset.

**Table S6 Significantly enriched gene sets genes identified in the SLE-MG-RA meta-analysis**.

The p.Val column is the adjusted p-value using the suggested g:SCS method by the g:Prolifer tool used for the analysis. The Genes column contains the provided genes that are in each identified gene set.

**Table S7 Genome-wide significant (p<5×10-8) LD independent loci from the T1D-MG-PSC meta-analysis**.

The column SNP contains the top SNP in each locus. The columns P, OR and SE correspond to the top SNP in each locus. The autoimmune disorder specific Top SNP column contains the top genome-wide significant SNP in the locus that was available in the input dataset. The autoimmune disorder specific P column contains the lowest p-value in the locus that was available in the input dataset.

**Table S8 Significantly enriched gene sets genes identified in the T1D-MG-PSC meta-analysis**.

The p.Val column is the adjusted p-value using the suggested g:SCS method by the g:Prolifer tool used for the analysis. The Genes column contains the provided genes that are in each identified gene set.

**Table S9 Genome-wide significant (p<5×10-8) LD independent loci from the PSC-MS meta-analysis**.

The column SNP contains the top SNP in each locus. The columns P, OR and SE correspond to the top SNP in each locus. The autoimmune disorder specific Top SNP column contains the top genome-wide significant SNP in the locus that was available in the input dataset. The autoimmune disorder specific P column contains the lowest p-value in the locus that was available in the input dataset.

**Table S10 Significantly enriched gene sets genes identified in the PSC-MS meta-analysis**. The p.Val column is the adjusted p-value using the suggested g:SCS method by the g:Prolifer tool used for the analysis. The Genes column contains the provided genes that are in each identified gene set.

**Figure S1 Overview of the phenotypes included in the general UK Biobank categories**. The number inside the circles shows the total number of phenotypes in each category. The numbers in each block show the percentage of phenotypes included in each sub-category.

**Figure S2 Overview of the phenotypes included in the general UK Biobank categories**. The number inside the circles shows the total number of phenotypes in each category. The numbers in each block show the percentage of phenotypes included in each sub-category

**Figure S3 Volcano plot of all PheWAS results for each autoimmune disorder**.

Annotated are the top 3 significant results, and with red the outcomes with p_FDR_<0.05.

**Figure S4 Significant PRS-PheWAS for at least three AD PRS with phenotypes in the Health and Medical History UK Biobank category**.

The shown phenotypes were significantly associated, after FDR adjustment, with at least three AD PRS irrespectively of the HLA status. The colors of cells indicate the standardized effect sizes (*β*) for the regression between AD PRS with HLA and each phenotype. The one star “☆” shows the significant results only with the “HLA included” AD PRS. The two stars “☆☆” show the significant associations with both “HLA included or excluded” AD PRS with the same effect direction. The star and the upper facing triangle “☆▵” show the significant associations with both “HLA included or excluded” AD PRS but with opposite effect directions. The upper facing triangle “▵” shows the significant associations only with “HLA excluded” AD PRS that the effect direction is the same as the color indicates. For the grouping of the phenotypes, we used the categories provided by the UK Biobank. We used the hclust R function to perform the hierarchical clustering of the autoimmune disorders showing in the dendrogram using all standardized effect sizes for the health and medical history category phenotypes.

**Figure S5 Significant PRS-PheWAS for at least three AD PRS with phenotypes in the Sociodemographics UK Biobank category**.

The shown phenotypes were significantly associated, after FDR adjustment, with at least three AD PRS irrespectively of the HLA status. The colors of cells indicate the standardized effect sizes (*β*) for the regression between AD PRS with HLA and each phenotype. The one star “☆” shows the significant results only with the “HLA included” AD PRS. The two stars “☆☆” show the significant associations with both “HLA included or excluded” AD PRS with the same effect direction. The star and the upper facing triangle “☆▵” show the significant associations with both “HLA included or excluded” AD PRS but with opposite effect directions. The upper facing triangle “▵” shows the significant associations only with “HLA excluded” AD PRS that the effect direction is the same as the color indicates. For the grouping of the phenotypes, we used the categories provided by the UK Biobank. We used the hclust R function to perform the hierarchical clustering of the autoimmune disorders showing in the dendrogram using all standardized effect sizes for the Sociodemographics category phenotypes.

**Figure S6 Significant PRS-PheWAS for at least three AD PRS with phenotypes in the Biomarkers (Blood Count) UK Biobank category**.

The shown phenotypes were significantly associated, after FDR adjustment, with at least three AD PRS irrespectively of the HLA status. The colors of cells indicate the standardized effect sizes (*β*) for the regression between AD PRS with HLA and each phenotype. The one star “☆” shows the significant results only with the “HLA included” AD PRS. The two stars “☆☆” show the significant associations with both “HLA included or excluded” AD PRS with the same effect direction. The star and the upper facing triangle “☆▵” show the significant associations with both “HLA included or excluded” AD PRS but with opposite effect directions. The upper facing triangle “▵” shows the significant associations only with “HLA excluded” AD PRS that the effect direction is the same as the color indicates. For the grouping of the phenotypes, we used the categories provided by the UK Biobank. We used the hclust R function to perform the hierarchical clustering of the autoimmune disorders showing in the dendrogram using all standardized effect sizes for the Biomarkers category phenotypes.

**Figure S7 Significant PRS-PheWAS for at least three AD PRS with phenotypes in the Biomarkers (Blood biochemistry) UK Biobank category**.

The shown phenotypes were significantly associated, after FDR adjustment, with at least three AD PRS irrespectively of the HLA status. The colors of cells indicate the standardized effect sizes (*β*) for the regression between AD PRS with HLA and each phenotype. The one star “☆” shows the significant results only with the “HLA included” AD PRS. The two stars “☆☆” show the significant associations with both “HLA included or excluded” AD PRS with the same effect direction. The star and the upper facing triangle “☆▵” show the significant associations with both “HLA included or excluded” AD PRS but with opposite effect directions. The upper facing triangle “▵” shows the significant associations only with “HLA excluded” AD PRS that the effect direction is the same as the color indicates. For the grouping of the phenotypes, we used the categories provided by the UK Biobank. We used the hclust R function to perform the hierarchical clustering of the autoimmune disorders showing in the dendrogram using all standardized effect sizes for the Biomarkers category phenotypes.

**Figure S8 Significant PRS-PheWAS for at least three AD PRS with phenotypes in the Biomarkers (Infectious Diseases) UK Biobank category**.

The shown phenotypes were significantly associated, after FDR adjustment, with at least three AD PRS irrespectively of the HLA status. The colors of cells indicate the standardized effect sizes (*β*) for the regression between AD PRS with HLA and each phenotype. The one star “☆” shows the significant results only with the “HLA included” AD PRS. For the grouping of the phenotypes, we used the categories provided by the UK Biobank. We used the hclust R function to perform the hierarchical clustering of the autoimmune disorders showing in the dendrogram using all standardized effect sizes for the Biomarkers category phenotypes.

**Figure S9 Significant PRS-PheWAS for at least three AD PRS with phenotypes in the Physical Measures UK Biobank category**.

The shown phenotypes were significantly associated, after FDR adjustment, with at least three AD PRS irrespectively of the HLA status. The colors of cells indicate the standardized effect sizes (*β*) for the regression between AD PRS with HLA and each phenotype. The one star “☆” shows the significant results only with the “HLA included” AD PRS. The two stars “☆☆” show the significant associations with both “HLA included or excluded” AD PRS with the same effect direction. The star and the upper facing triangle “☆▵” show the significant associations with both “HLA included or excluded” AD PRS but with opposite effect directions. The upper facing triangle “▵” shows the significant associations only with “HLA excluded” AD PRS that the effect direction is the same as the color indicates. For the grouping of the phenotypes, we used the categories provided by the UK Biobank. We used the hclust R function to perform the hierarchical clustering of the autoimmune disorders showing in the dendrogram using all standardized effect sizes for the Physical Measures category phenotypes.

**Figure S10 Genetic correlation of the 11 autoimmune disorders**

The figure shows the genetic correlation for all 11 autoimmune disorders included in the analysis. A) After excluding the SNPs in the HLA region (hg19, chr6 25-33 Mb). B) Without excluding the SNPs in the HLA region. The red color in the cells reflects more positive correlation coefficients while blue reflects more negative coefficients, and the numbers within each cell are the correlation coefficients. The correlations with p<0.05 are denoted with one asterisk (*), while the two asterisks show the correlations that are significant after the Bonferroni correction.

