## Supplementary figures for "PheWAS and cross-disorder analyses reveal genetic architecture, pleiotropic loci and phenotypic correlations across 11 autoimmune disorders"

Topaloudi et al.

Figure S1 Overview of the phenotypes included in the general UK Biobank categories. 3

Figure S2 Overview of the phenotypes included in the general UK Biobank categories. 4

Figure S3 Volcano plot of all PheWAS results for each autoimmune disorder. 9

Figure S4 Significant PRS-PheWAS for at least three AD PRS with phenotypes in the Health and Medical History UK Biobank category. 10

Figure S5 Significant PRS-PheWAS for at least three AD PRS with phenotypes in the Sociodemographics UK Biobank category. 12

Figure S6 Significant PRS-PheWAS for at least three AD PRS with phenotypes in the Biomarkers (Blood Count) UK Biobank category. 14

Figure S7 Significant PRS-PheWAS for at least three AD PRS with phenotypes in the Biomarkers (Blood biochemistry) UK Biobank category. 16

Figure S8 Significant PRS-PheWAS for at least three AD PRS with phenotypes in the Biomarkers (Infectious Diseases) UK Biobank category. 17

Figure S9 Significant PRS-PheWAS for at least three AD PRS with phenotypes in the Physical Measures UK Biobank category. 18

Figure S10 Genetic correlation of the 11 autoimmune disorders. 19

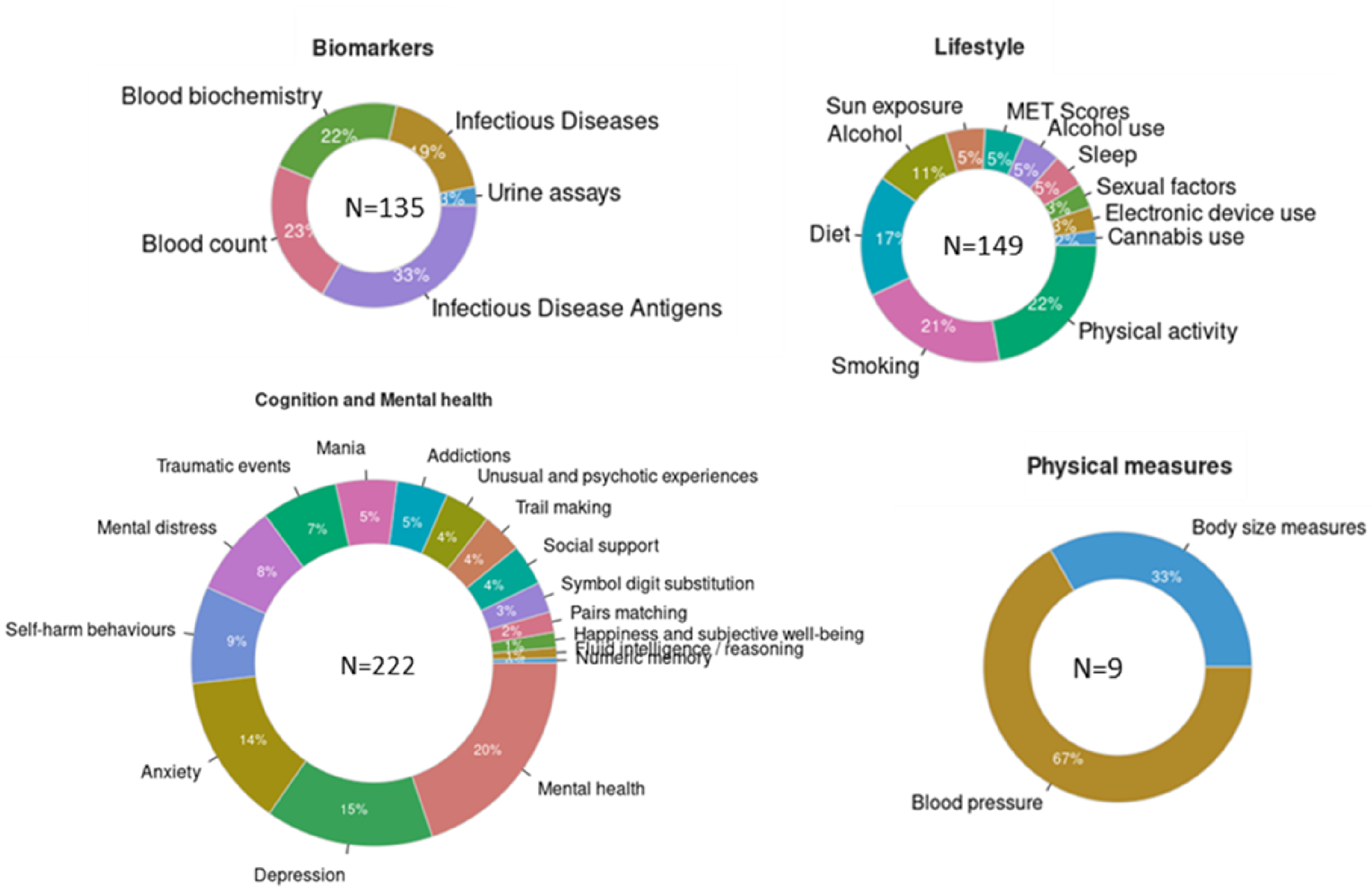


**Figure S1 Overview of the phenotypes included in the general UK Biobank categories.**
The number inside the circles shows the total number of phenotypes in each category. The numbers in each block show the percentage of phenotypes included in each sub-category.

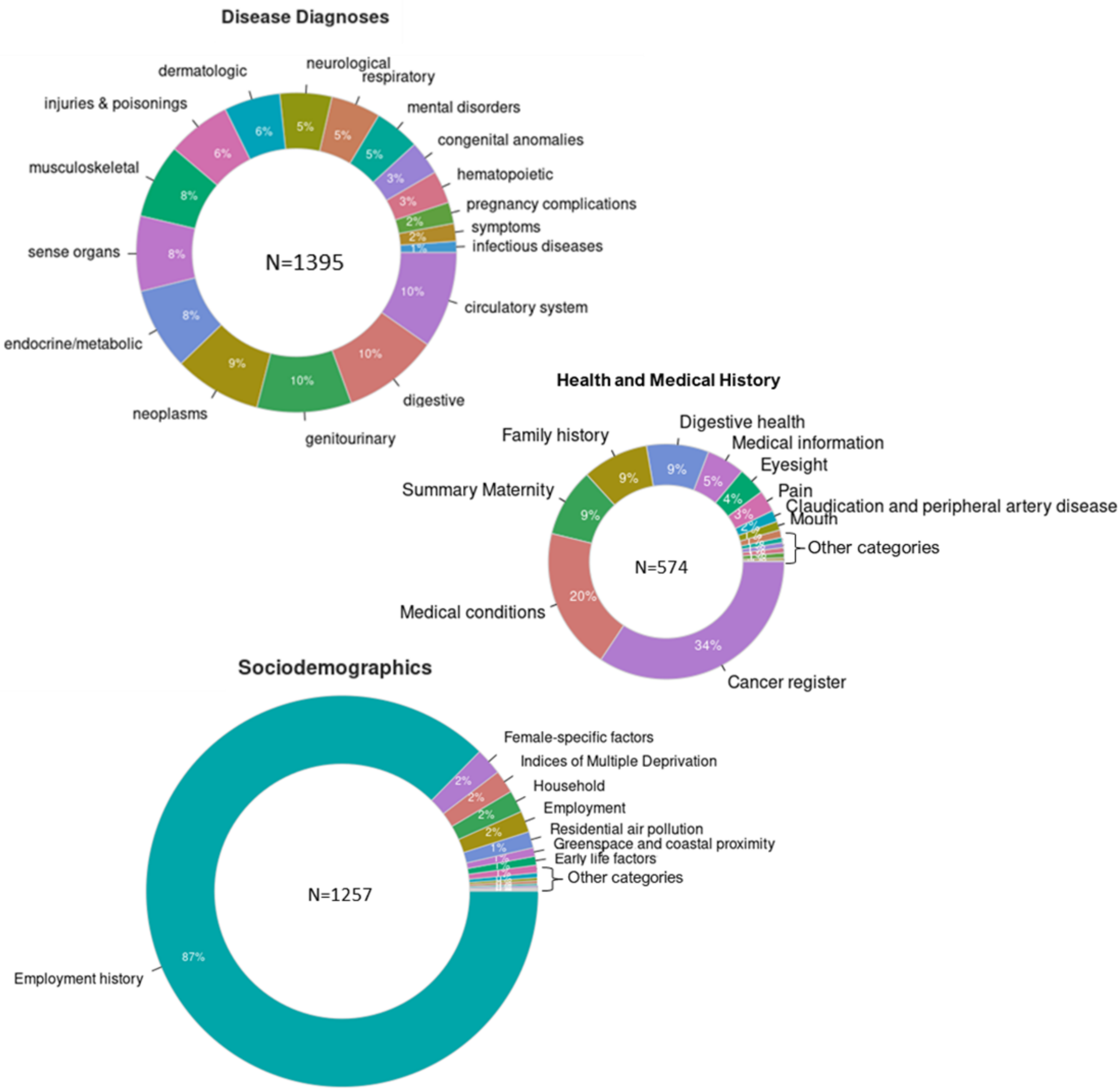


**Figure S2 Overview of the phenotypes included in the general UK Biobank categories.** The number inside the circles shows the total number of phenotypes in each category. The numbers in each block show the percentage of phenotypes included in each sub-category.

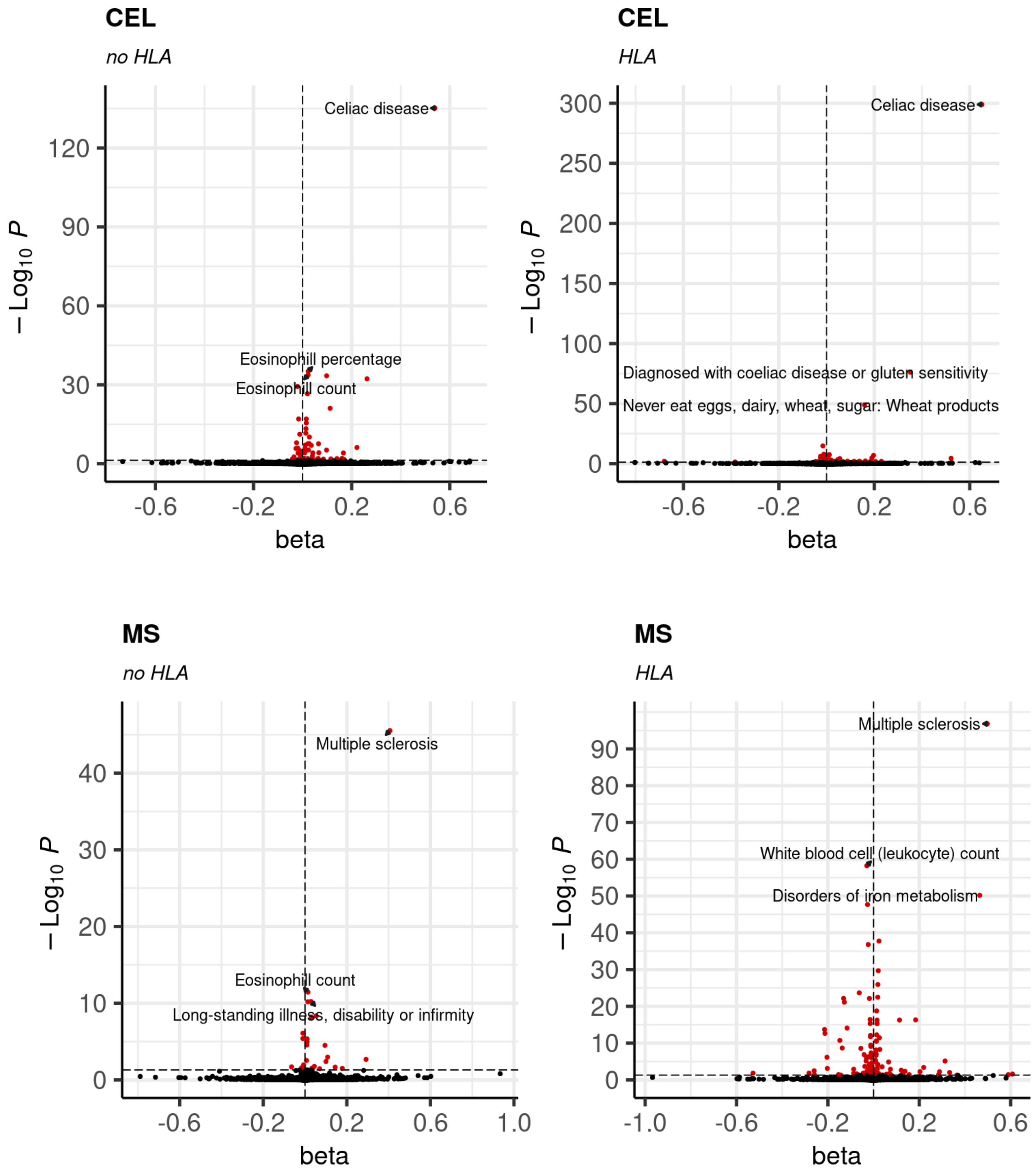
CEL
no HLA
Celiac disease
Eosinophill percentage
Eosinophill count
120
90
60
30
0
-0.6
-0.2
0.2
0.6
beta
$-\log_{10} P$
CEL
HLA
Celiac disease
Diagnosed with coeliac disease or gluten sensitivity
Never eat eggs, dairy, wheat, sugar: Wheat products
300
250
200
150
100
50
0
-0.6
-0.2
0.2
0.6
beta
$-\log_{10} P$
MS
no HLA
Multiple sclerosis
Eosinophill count
Long-standing illness, disability or infirmity
40
30
20
10
0
-0.6
-0.2
0.2
0.6
1.0
beta
$-\log_{10} P$
MS
HLA
Multiple sclerosis
White blood cell (leukocyte) count
Disorders of iron metabolism
90
80
70
60
50
40
30
20
10
0
-1.0
-0.6
-0.2
0.2
0.6
beta
$-\log_{10} P$

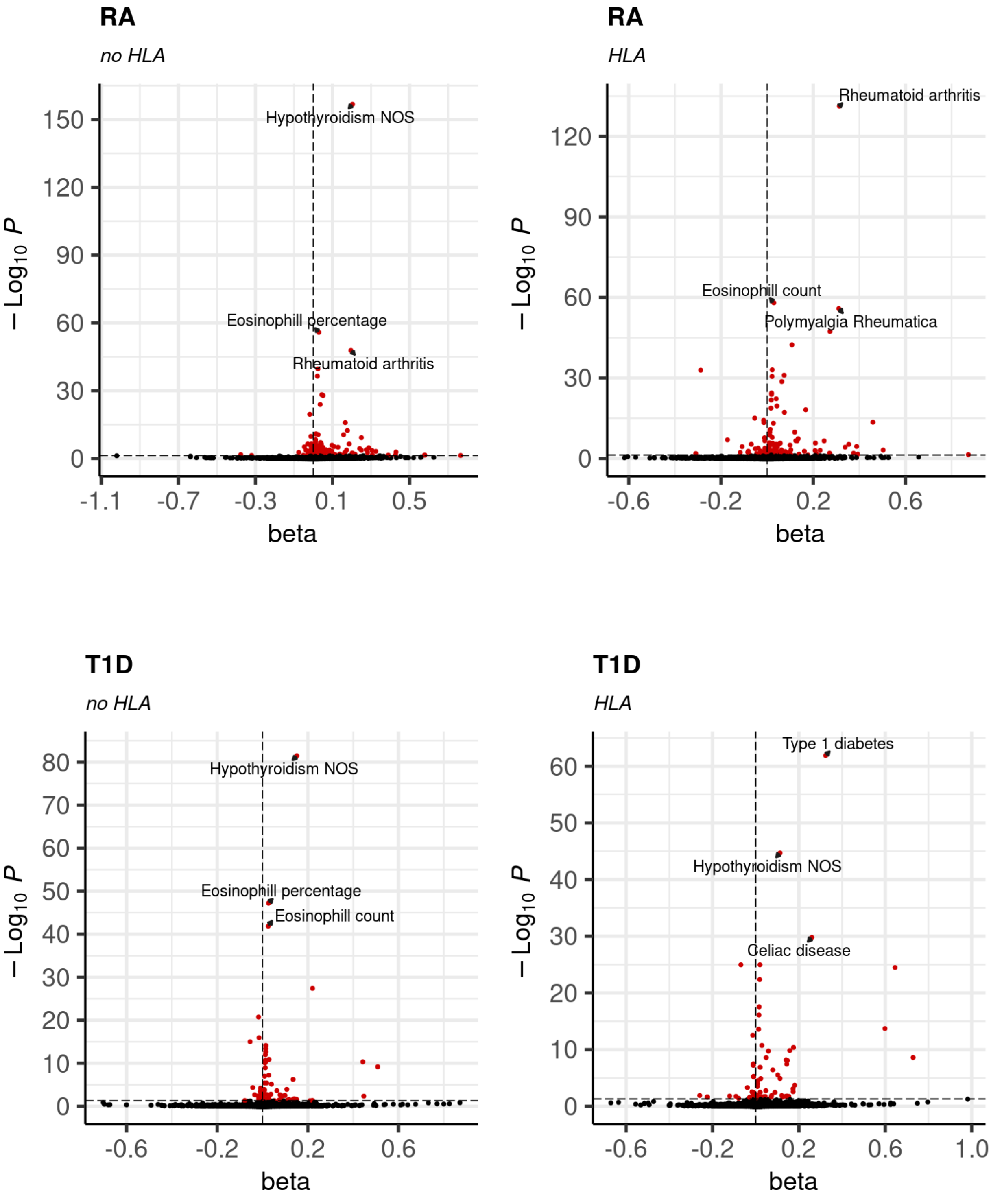
RA
no HLA
Hypothyroidism NOS
Eosinophill percentage
Rheumatoid arthritis
150
120
90
60
30
0
-1.1
-0.7
-0.3
0.1
0.5
beta
$-\log_{10} P$
RA
HLA
Rheumatoid arthritis
Eosinophill count
Polymyalgia Rheumatica
120
90
60
30
0
-0.6
-0.2
0.2
0.6
beta
$-\log_{10} P$
T1D
no HLA
Hypothyroidism NOS
Eosinophill percentage
Eosinophill count
80
70
60
50
40
30
20
10
0
-0.6
-0.2
0.2
0.6
beta
$-\log_{10} P$
T1D
HLA
Type 1 diabetes
Hypothyroidism NOS
Celiac disease
60
50
40
30
20
10
0
-0.6
-0.2
0.2
0.6
1.0
beta
$-\log_{10} P$

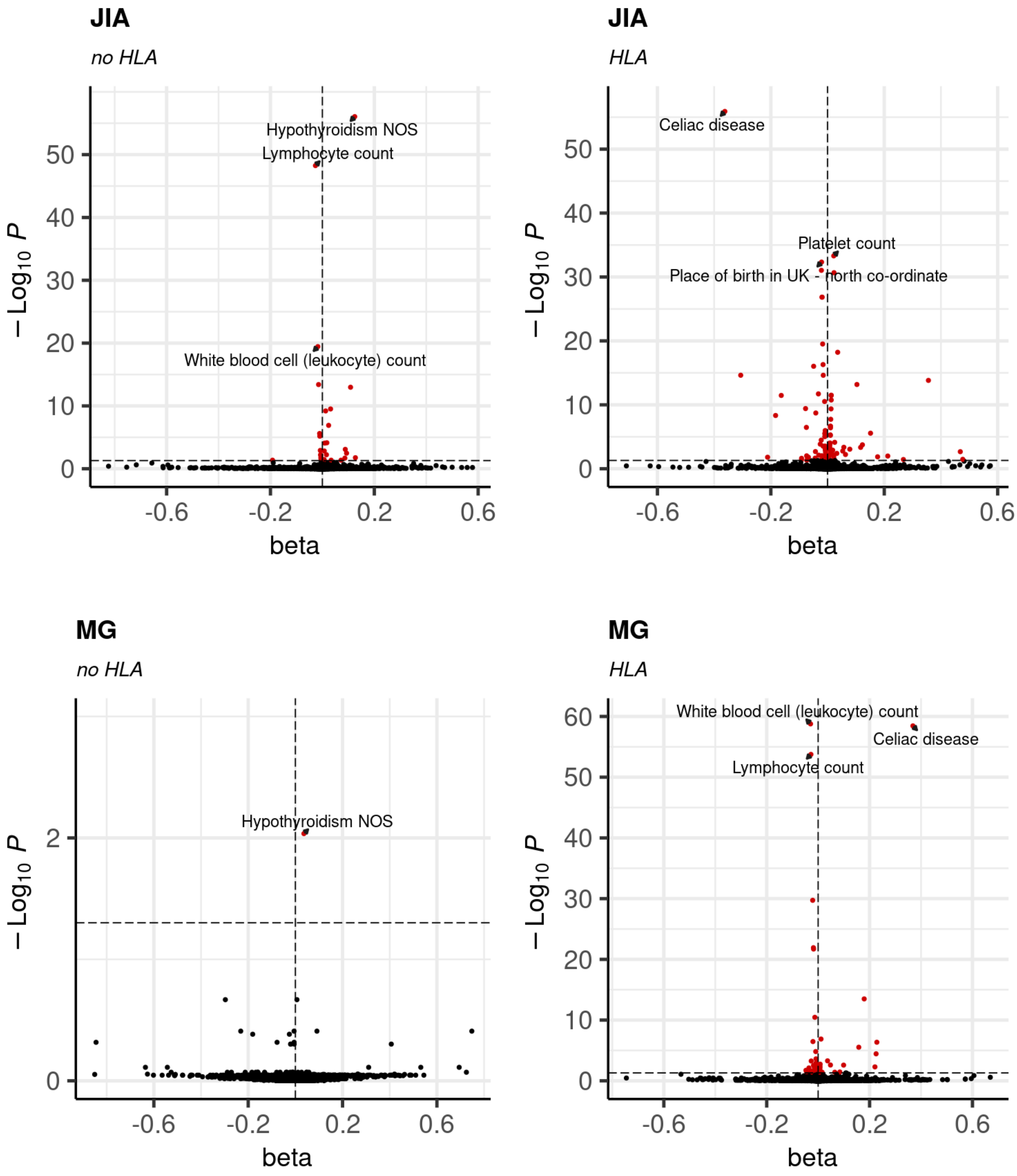
JIA
no HLA
Hypothyroidism NOS
Lymphocyte count
White blood cell (leukocyte) count
JIA
HLA
Celiac disease
Platelet count
Place of birth in UK - north co-ordinate
MG
no HLA
Hypothyroidism NOS
MG
HLA
White blood cell (leukocyte) count
Celiac disease
Lymphocyte count
beta
$-\text{Log}_{10}\,P$

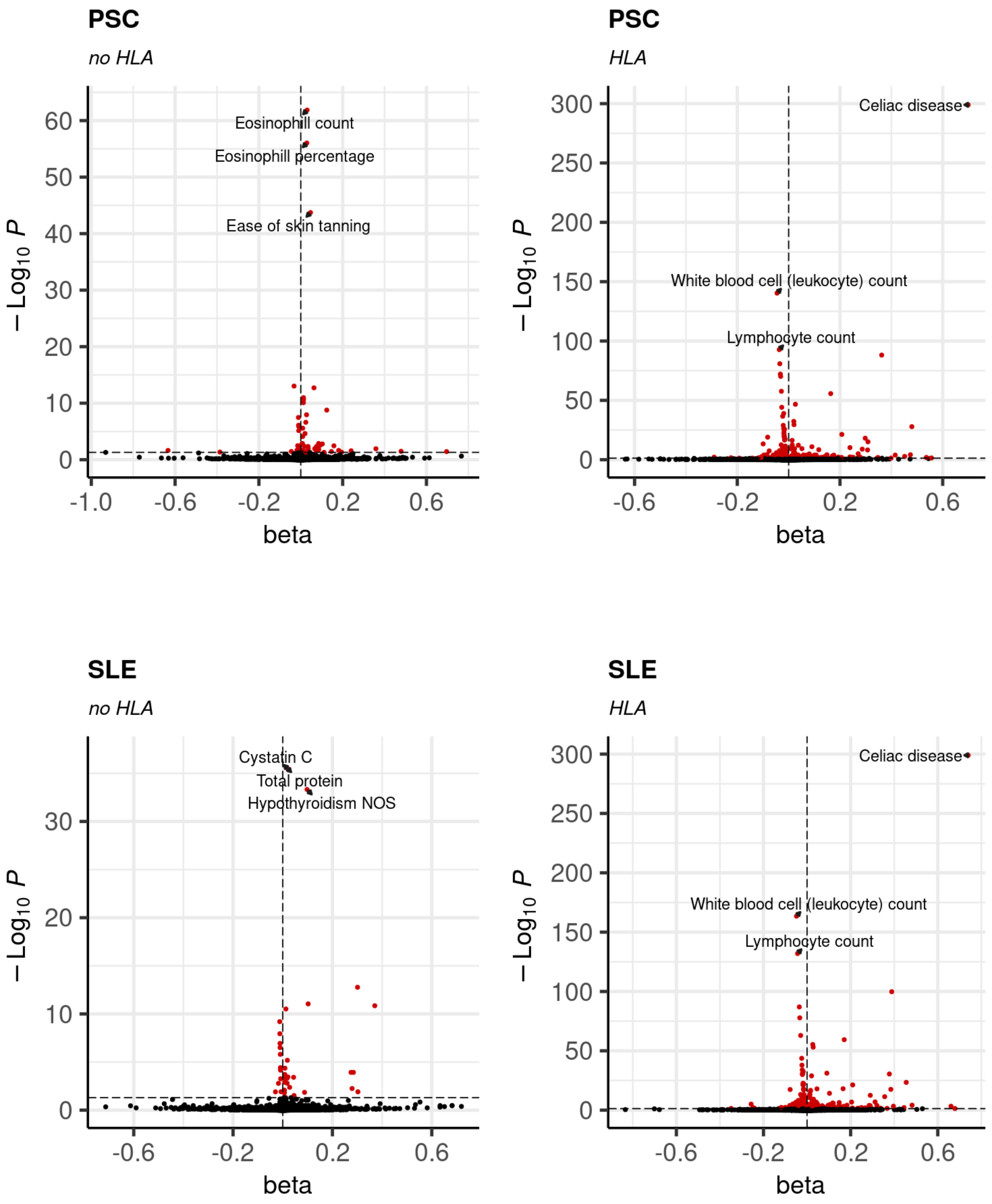
PSC
no HLA
Eosinophill count
Eosinophill percentage
Ease of skin tanning
$-\text{Log}_{10}\ P$
beta
PSC
HLA
Celiac disease
White blood cell (leukocyte) count
Lymphocyte count
$-\text{Log}_{10}\ P$
beta
SLE
no HLA
Cystatin C
Total protein
Hypothyroidism NOS
$-\text{Log}_{10}\ P$
beta
SLE
HLA
Celiac disease
White blood cell (leukocyte) count
Lymphocyte count
$-\text{Log}_{10}\ P$
beta

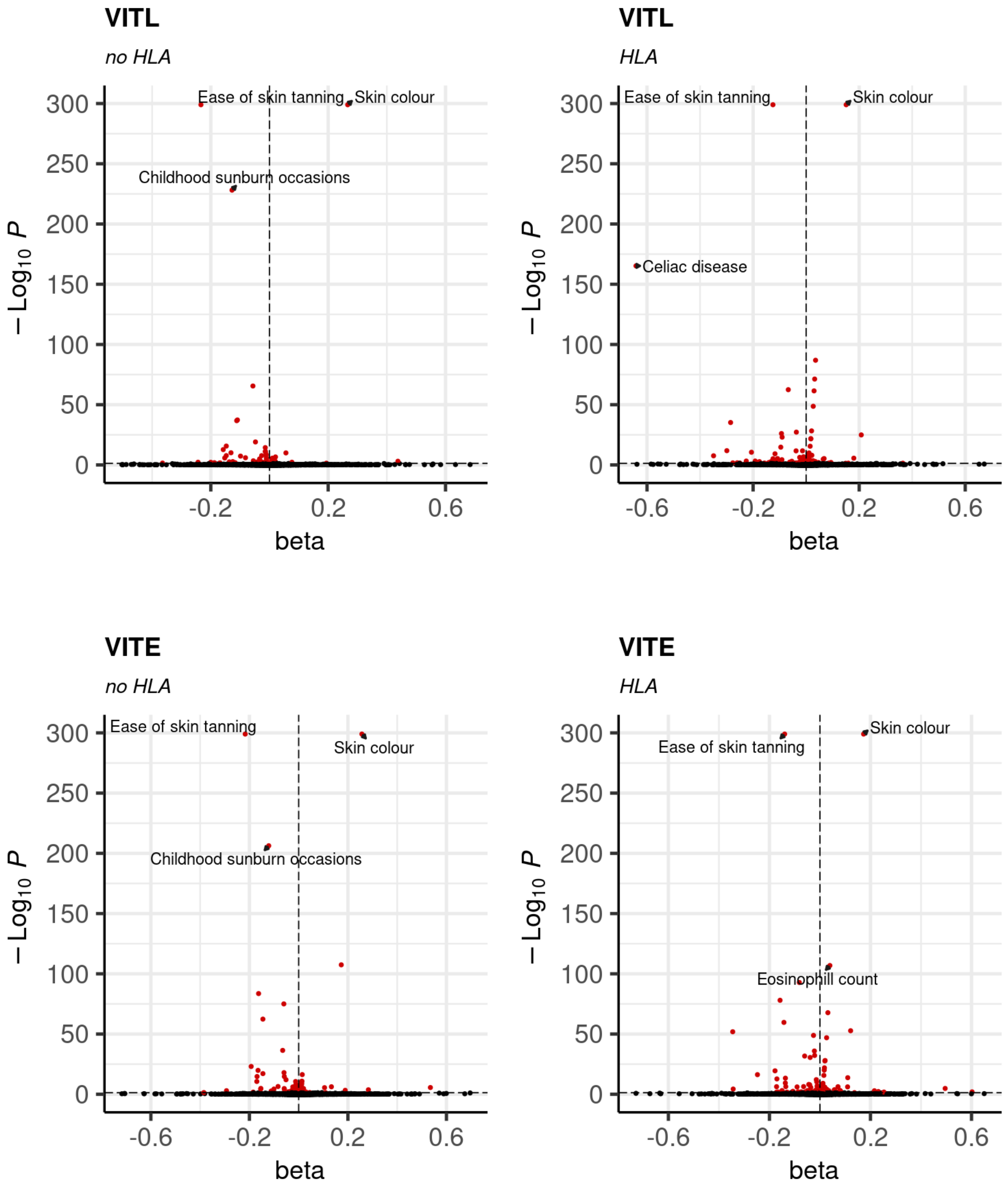


**Figure S3 Volcano plot of all PheWAS results for each autoimmune disorder.**
Annotated are the top 3 significant results, and with red the outcomes with pFDR<0.05.

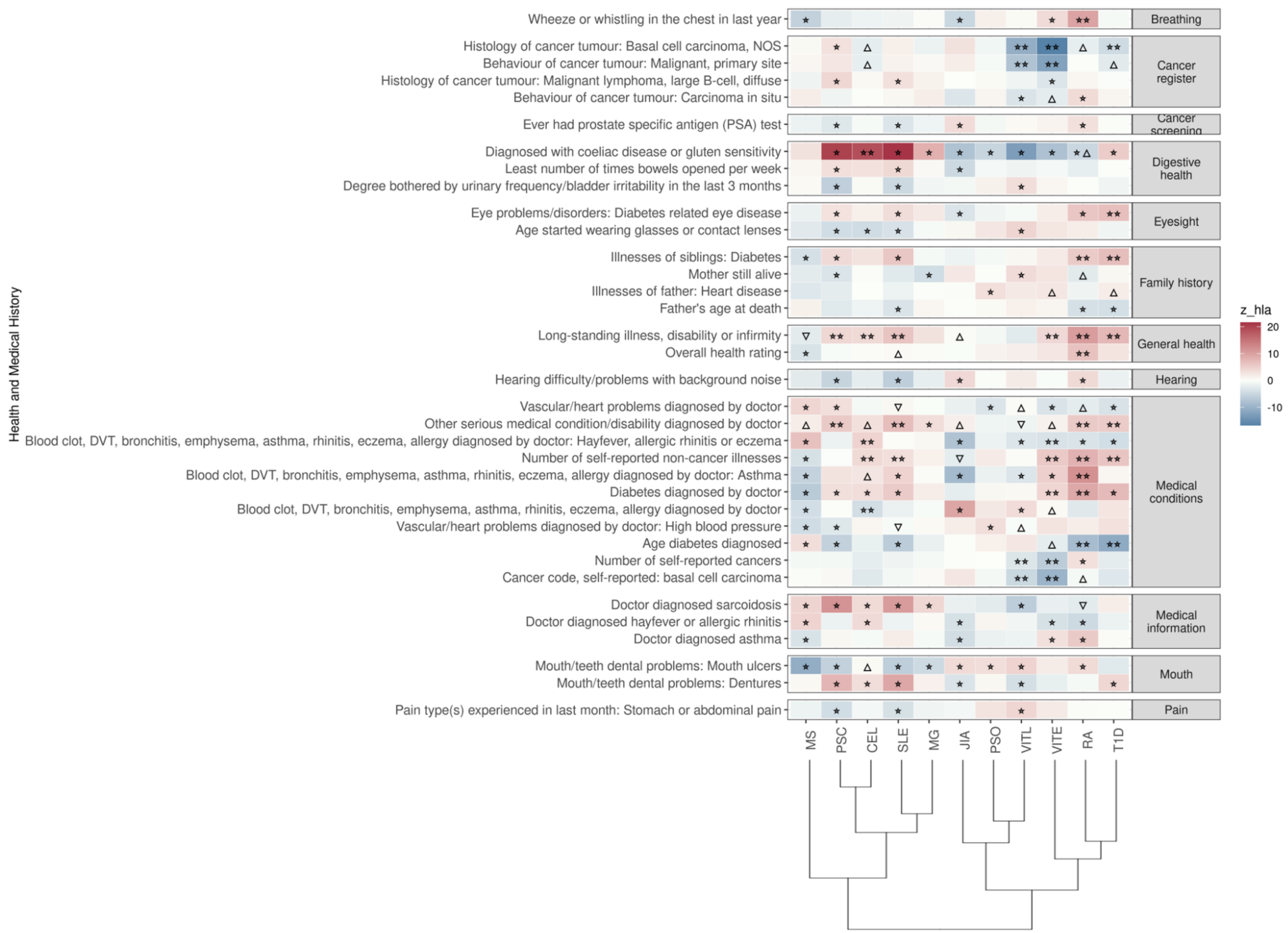


**Figure S4 Significant PRS-PheWAS for at least three AD PRS with phenotypes in the Health and Medical History UK Biobank category.** The shown phenotypes were significantly associated, after FDR adjustment, with at least three AD PRS irrespectively of the HLA status. The colors of cells indicate the standardized effect sizes (β) for the regression between AD PRS with HLA and each phenotype. The one star "☆"

shows the significant results only with the “HLA included” AD PRS. The two stars “☆☆” show the significant associations with both “HLA included or excluded” AD PRS with the same effect direction. The star and the upper facing triangle “☆▵” show the significant associations with both “HLA included or excluded” AD PRS but with opposite effect directions. The upper facing triangle “▵” shows the significant associations only with “HLA excluded” AD PRS that the effect direction is the same as the color indicates. For the grouping of the phenotypes, we used the categories provided by the UK Biobank. We used the hclust R function to perform the hierarchical clustering of the autoimmune disorders showing in the dendrogram using all standardized effect sizes for the health and medical history category phenotypes.

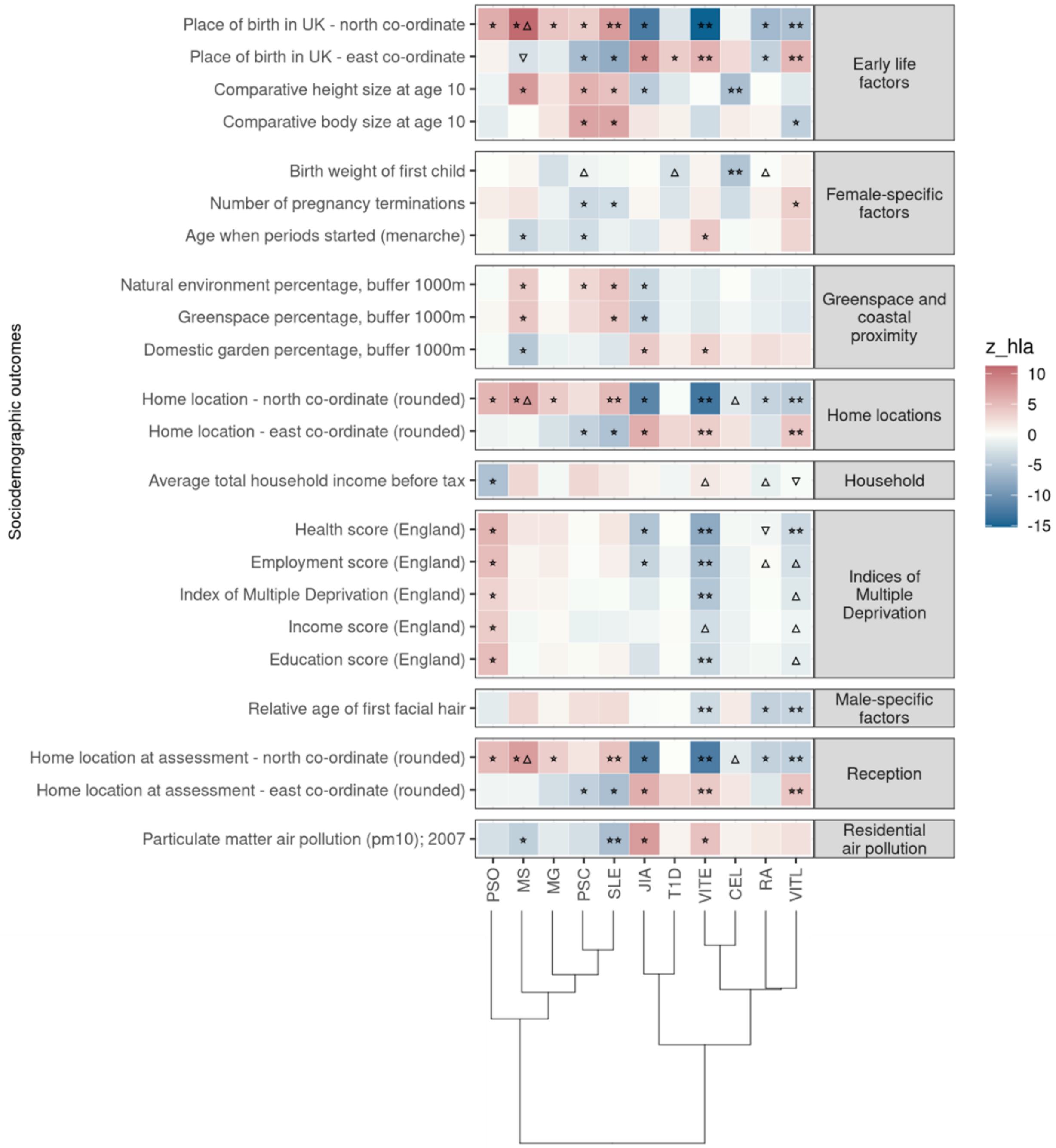


**Figure S5 Significant PRS-PheWAS for at least three AD PRS with phenotypes in the Sociodemographics UK Biobank category.**

The shown phenotypes were significantly associated, after FDR adjustment, with at least three AD PRS irrespectively of the HLA status. The colors of cells indicate the standardized effect sizes (β) for the regression between AD PRS with HLA and each phenotype. The one star "☆" shows the significant results only with the "HLA included" AD PRS. The two stars "☆☆" show the significant associations with both "HLA included or excluded" AD PRS with the same effect direction. The star and the upper facing triangle “☆▵” show the significant associations with both “HLA included or excluded” AD PRS but with opposite effect directions. The upper facing triangle “▵” shows the significant associations only with “HLA excluded” AD PRS that the effect direction is the same as the color indicates. For the grouping of the phenotypes, we used the categories provided by the UK Biobank. We used the hclust R function to perform the hierarchical clustering of the autoimmune disorders showing in the dendrogram using all standardized effect sizes for the Sociodemographics category phenotypes.

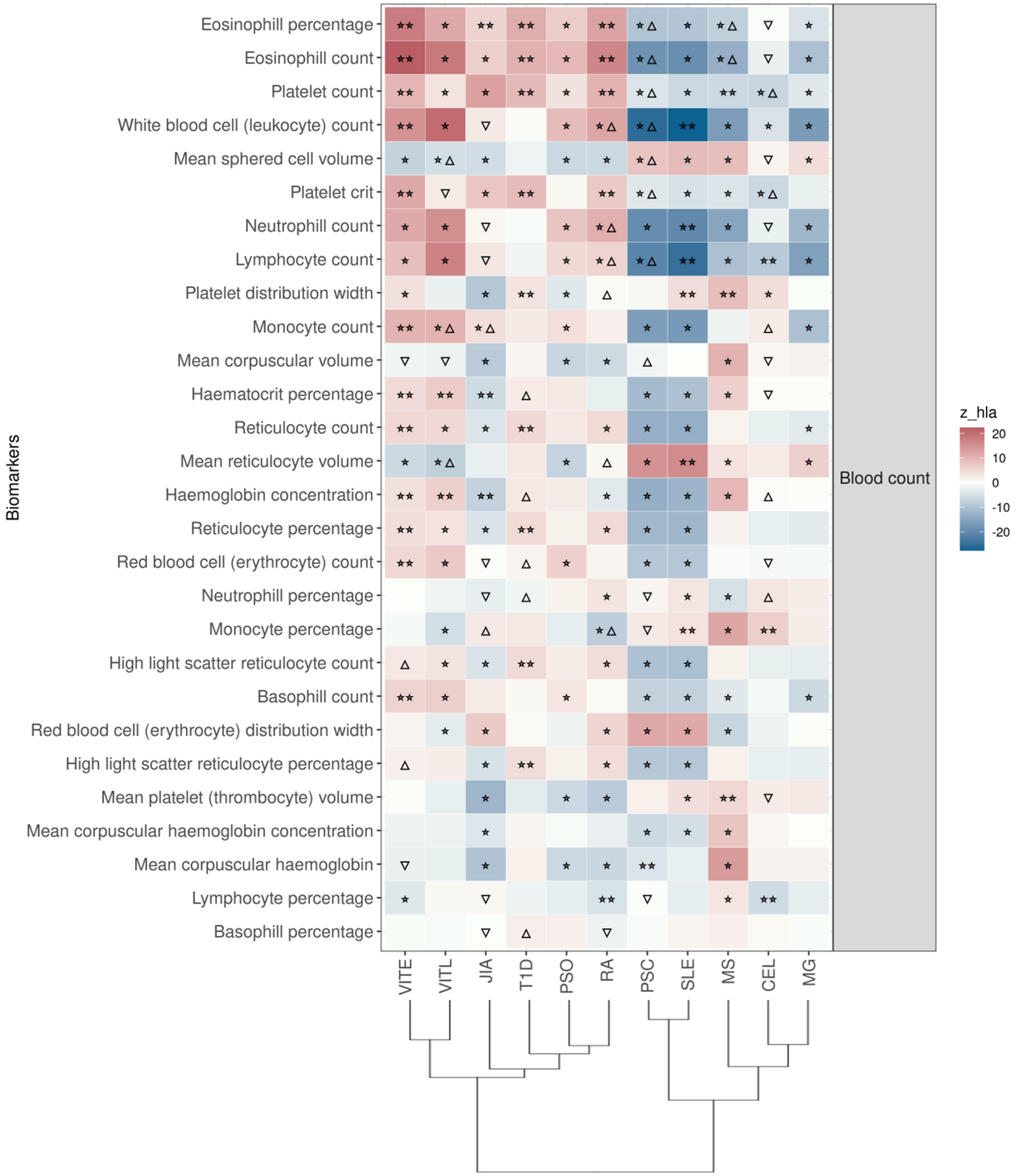


**Figure S6 Significant PRS-PheWAS for at least three AD PRS with phenotypes in the Biomarkers (Blood Count) UK Biobank category.**

The shown phenotypes were significantly associated, after FDR adjustment, with at least three AD PRS irrespectively of the HLA status. The colors of cells indicate the standardized effect sizes (β) for the regression between AD PRS with HLA and each phenotype. The one star "☆" shows the significant results only with the "HLA included" AD PRS. The two stars "☆☆" show the significant associations with both "HLA included or excluded" AD PRS with the same effect direction. The star and the upper facing triangle “☆▵” show the significant associations with both “HLA included or excluded” AD PRS but with opposite effect directions. The upper facing triangle “▵” shows the significant associations only with “HLA excluded” AD PRS that the effect direction is the same as the color indicates. For the grouping of the phenotypes, we used the categories provided by the UK Biobank. We used the hclust R function to perform the hierarchical clustering of the autoimmune disorders showing in the dendrogram using all standardized effect sizes for the Biomarkers category phenotypes.

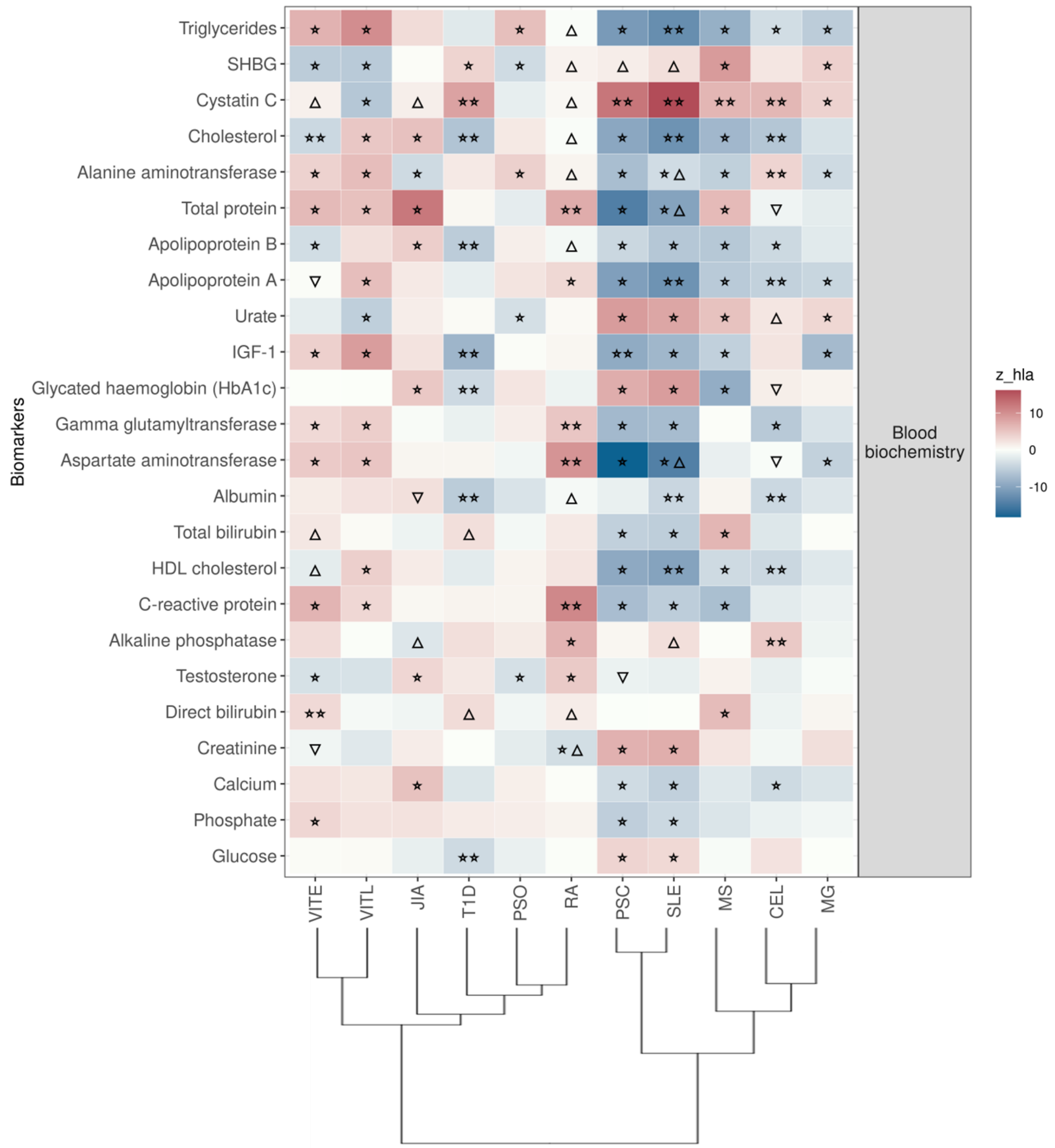


**Figure S7 Significant PRS-PheWAS for at least three AD PRS with phenotypes in the Biomarkers (Blood biochemistry) UK Biobank category.**

The shown phenotypes were significantly associated, after FDR adjustment, with at least three AD PRS irrespectively of the HLA status. The colors of cells indicate the standardized effect sizes (β) for the regression between AD PRS with HLA and each phenotype. The one star "☆" shows the significant results only with the "HLA included" AD PRS. The two stars "☆☆" show the significant associations with both "HLA included or excluded" AD PRS with the same effect direction. The star and the upper facing triangle "☆▵" show the significant associations with both "HLA included or excluded" AD PRS but with opposite effect directions. The upper facing triangle "▵" shows the significant associations only with “HLA excluded” AD PRS that the effect direction is the same as the color indicates. For the grouping of the phenotypes, we used the categories provided by the UK Biobank. We used the hclust R function to perform the hierarchical clustering of the autoimmune disorders showing in the dendrogram using all standardized effect sizes for the Biomarkers category phenotypes.

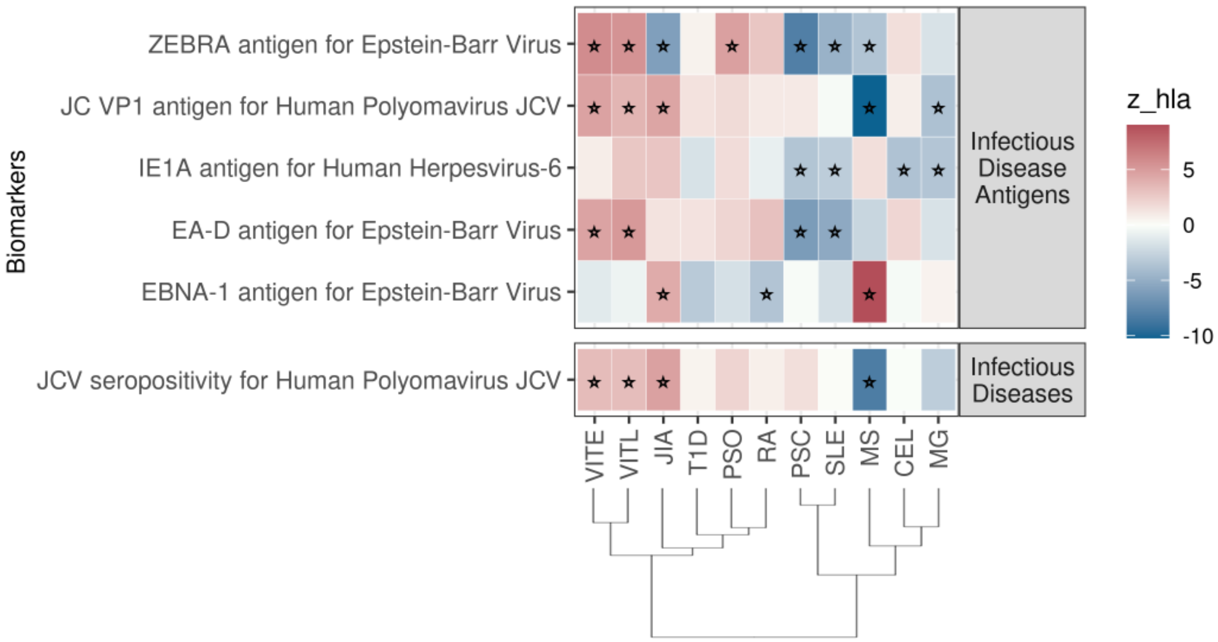


**Figure S8 Significant PRS-PheWAS for at least three AD PRS with phenotypes in the Biomarkers (Infectious Diseases) UK Biobank category.**

The shown phenotypes were significantly associated, after FDR adjustment, with at least three AD PRS irrespectively of the HLA status. The colors of cells indicate the standardized effect sizes (β) for the regression between AD PRS with HLA and each phenotype. The one star "☆" shows the significant results only with the "HLA included" AD PRS. For the grouping of the phenotypes, we used the categories provided by the UK Biobank. We used the hclust R function to perform the hierarchical clustering of the autoimmune disorders showing in the dendrogram using all standardized effect sizes for the Biomarkers category phenotypes.

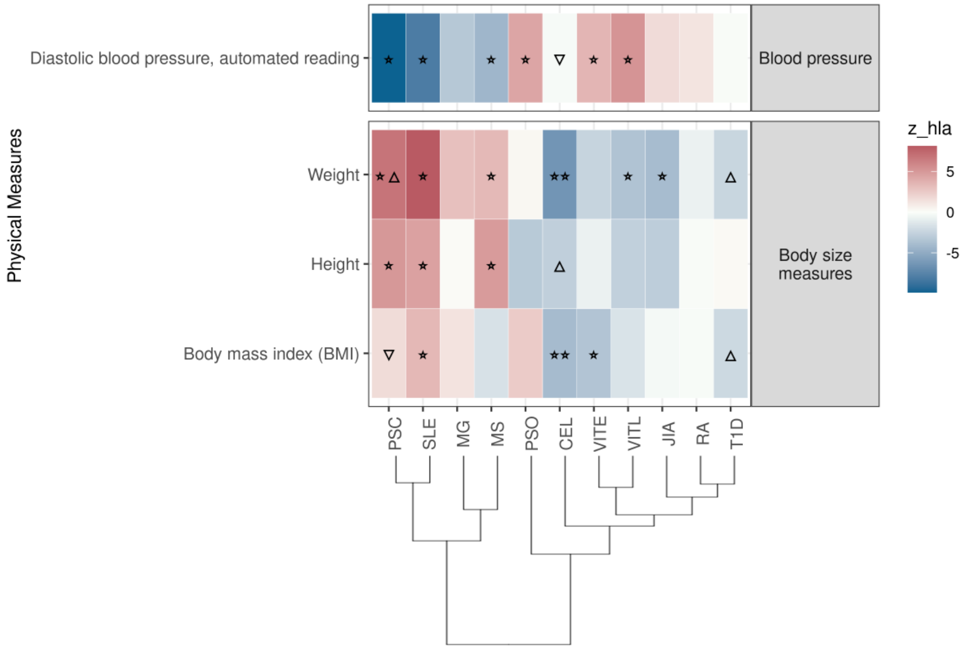


**Figure S9 Significant PRS-PheWAS for at least three AD PRS with phenotypes in the Physical Measures UK Biobank category.**

The shown phenotypes were significantly associated, after FDR adjustment, with at least three AD PRS irrespectively of the HLA status. The colors of cells indicate the standardized effect sizes (β) for the regression between AD PRS with HLA and each phenotype. The one star "☆" shows the significant results only with the "HLA included" AD PRS. The two stars "☆☆" show the significant associations with both "HLA included or excluded" AD PRS with the same effect direction. The star and the upper facing triangle "☆▵" show the significant associations with both "HLA included or excluded" AD PRS but with opposite effect directions. The upper facing triangle "▵" shows the significant associations only with "HLA excluded" AD PRS that the effect direction is the same as the color indicates. For the grouping of the phenotypes, we used the categories provided by the UK Biobank. We used the hclust R function to perform the hierarchical clustering of the autoimmune disorders showing in the dendrogram using all standardized effect sizes for the Physical Measures category phenotypes.

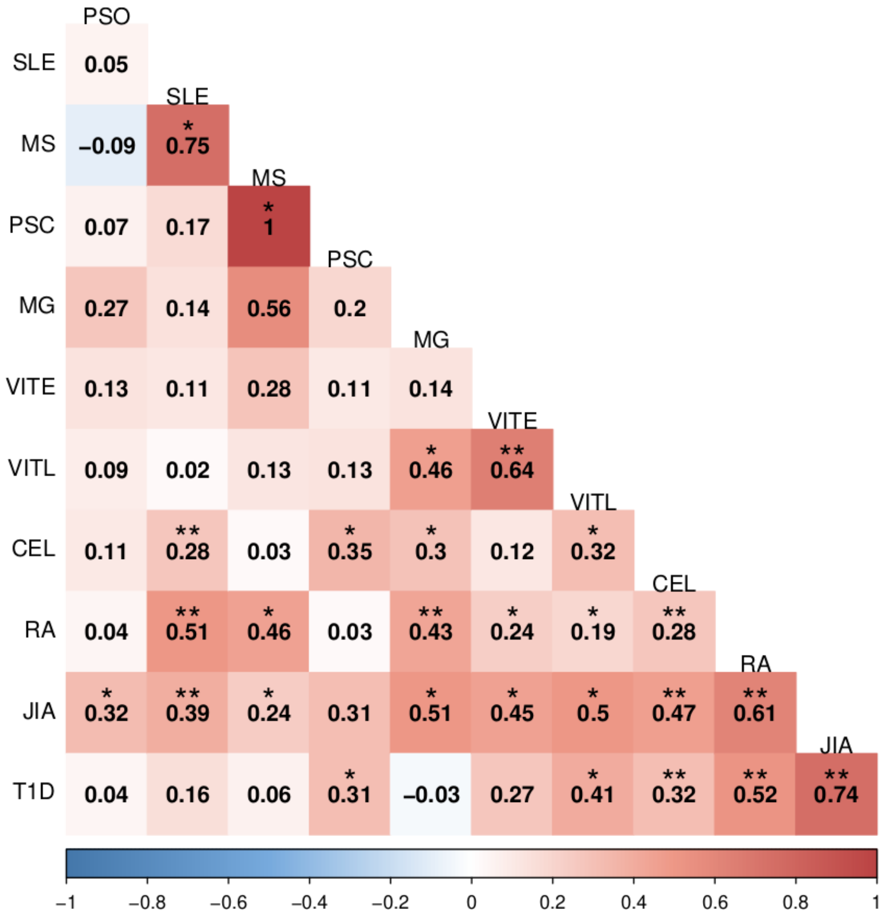


**Figure S10 Genetic correlation of the 11 autoimmune disorders.**

The figure shows the genetic correlation for all 11 autoimmune disorders included in the analysis. A) After excluding the SNPs in the HLA region (hg19, chr6 25-33 Mb). B) Without excluding the SNPs in the HLA region. The red color in the cells reflects more positive correlation coefficients while blue reflects more negative coefficients, and the numbers within each cell are the correlation coefficients. The correlations with p<0.05 are denoted with one asterisk (*), while the two asterisks show the correlations that are significant after the Bonferroni correction.
